# Flushing water regularly inadequate to protect water quality in gravity-fed water supply systems with storage tanks during long-term lockdown

**DOI:** 10.1101/2023.01.26.23284807

**Authors:** Deepika Bhaskar, Gargi Singh

## Abstract

Flushing of water is recommended to prevent the growth of opportunistic pathogens, corrosion, and deterioration of the water supply network during short-term lockdowns. However, the efficacy of flushing fixtures in water supply networks with intermittent gravity-fed supply during long-term lockdowns is unknown. A laboratory-scale premise water supply network with a secondary storage tank connected to a gravity-fed drinking water distribution network was operated for 52 weeks to compare the efficacy of flushing water once a day (1D) vs. once in three days (3D) to preserve water quality. The tap water sampled from buildings (building) and overhead tanks (OHTs) prior to the pandemic served as a benchmark for comparison. The water quality trended as 1D = 3D < building < OHTs, with both the 1D and 3D yielding ‘poor’ quality water. The water in 3D had higher levels of 16S rRNA gene copies, *sul*1, and *int*I1 relative to 1D; this difference was more pronounced (2-3 orders of magnitude) during summer. The levels of *sul*1, *int*I1, and DNA marker of *Legionella pneumophila* in biofilms sampled at the end of the operation of the laboratory-scale premise water supply network trended as elbows > visibly corroded pipes-sections > taps. The levels of heavy metals in some tap water samples exceeded the WHO recommendations and BIS standards for drinking water: 32% of 1D samples (30/93), and 31% of 3D samples (29/93) exceeded the standards for Pb, (10 ppb); while 48% (45/93) of 1D samples and 35% (33/93) of 3D samples exceeded the standards for Fe in drinking water (300 ppb).

## 1. Introduction

While regular flushing is advised to protect water quality during periods of reduced use or prolonged inactivity (Richard and Boyer, 2021), the efficacy of such guidelines for gravity-fed water supply systems that duplicate storage to cope with intermittent supply is unknown. In the water-scarce and -stressed countries (World Resources Institute, 2019), such as India, intermittent water supply and use of secondary water storage within premises are common (McGuinness et al., 2020; Shaheed et al., 2014). Even during regular use, gravity-fed overhead tank-based water supply systems are notorious for poor water quality (Bhaskar and Singh, 2021; Cookson and Stirk, 2019), increased corrosion (Li et al., 2016) and microbial growth (Douterelo et al., 2016b). How these already challenged gravity-fed, secondary storage tank-based water networks can be protected during prolonged building-specific lockdowns, as seen in the first two years of the COVID-19 pandemic, is unknown. The issues around the gravity-fed water supply networks with intermittent water supply may be limited mainly to third-world countries; however, they affect the health of billions of people globally (Bhaskar and Singh, 2021). Hence, it is imperative to comprehensively investigate the efficacy of strategies to protect against the deterioration of water supply networks (corrosion) and the growth of opportunistic pathogens during long-term lockdowns.

The escalation of water stagnation in intermittent and gravity-fed water supply systems at multiple levels (municipal and premise) provides more opportunities for corrosion, disinfectant decay, and growth of microbes, including opportunistic pathogens. Consequently, the interventions that work for fully pressurized continuous water supply systems during no-or reduced-use periods may be less helpful for gravity-fed intermittent water supply systems with premise-based water storage tanks. In the target region (north India), the premise-based secondary water storage tanks, referred to hereafter as ‘secondary storage tanks’, are typically PVC or RCC-based 500-1000 L water tanks, which directly receive either the underground water or municipally supplied water before supplying to the premises via gravity. These premise-based secondary storage tanks are typically neither cleaned nor disinfected regularly and are susceptible to microbial growth and contamination (Cookson and Stirk, 2019). On the community level, the (primary) overhead tanks (OHTs) and in the premises, the secondary water storage tanks are associated with compromised longevity of the water supply system, increased water age, and duplicated storage (Bhaskar and Singh, 2021). Even if safe water is supplied through the piped network, water quality can deteriorate within the premise-based storage tanks, especially in intensely contaminated environments (McGuinness et al., 2020). Nevertheless, the demand for water storage tanks continues to grow due to ever-increasing populations and interruptions to the water supply.

Generally, such systems also use chlorine-based disinfectants that are linked with increased antibiotic resistance potential (Bhaskar and Singh, 2021). Additionally, even during regular use, the overhead tank-based water supply systems tend to have a high prevalence of opportunistic pathogens and select for class 1 integron-integrase gene cassette (Bhaskar and Singh, 2021).

The efficacy of regularly flushing water during reduced use to protect water quality is debatable even in fully-pressurized and continuous water supply systems. While the poor-quality water can be flushed out entirely be before resuming regular use of the premises after the lockdown, any improvement in water quality may not last long (Hozalski et al., 2020). The colonization of biofilms by opportunistic pathogens and any deterioration of the integrity of the water supply network during the reduced or no-use periods could continue to affect the water safety post-flush (Ye et al., 2022). Short-term investigations (1 month) in locked-down residential buildings with continuous and fully-pressurised water supply have only shown a temporary benefit of flushing the water regularly, followed by the rapid deterioration of water quality post-flushing, indicating possible impact of compromised water network due to corrosion or excessive growth of biofilms during the lockdown (Hozalski et al., 2020). Thus, long-term investigations in both biofilm and tap water are needed. The time scale of the COVID-19 pandemic further emphasizes the need to investigate the effects of long-term pandemic-related restrictions on the water supply systems.

We investigated 1) the impact of flushing water once a day (1D), and II) the effect of reducing the frequency at which the water was flushed to once in 3 days (3D) on the water quality, levels of antibiotic resistance gene markers (*sul*1, *sul*2, *erm*F, *bla*OXA-1 and *int*I1), total bacterial count and the DNA markers of opportunistic pathogens (*Legionella pneumophila* and *Mycobacterium avium*) over a building-specific year-long simulated lockdown. The laboratory-scale premise water supply network employed a 100 L PVC-based secondary storage tank, which received disinfected groundwater intermittently from an OHT-based water supply network that was under regular use throughout the study, with stagnation limited to the laboratory-scale premise water supply network. Such building-specific stagnation was common during the pandemic-related lockdowns in 2020 in India and elsewhere (McGuinness et al., 2020). The first flush from the laboratory scale premise plumbing network was collected every twelfth day, and the biofilms were swabbed after the end of its year-long operation. Water conductivity, pH, total organic carbon, nitrate, sulfate, chloride, and heavy metals (Fe, Pb, Zn, and Cu) were measured in tap water. The water quality, prevalence of the opportunistic pathogens, and antibiotic resistance gene markers in tap water under 1D and 3D were compared with pre-pandemic building and OHT water samples. Throughout the experiment, the water quality of the first flush deteriorated under both 1D and 3D approaches, with increase in the prevalence of the DNA marker of *L. pneumophila* and the proportion of samples that failed to meet the drinking water standards for Fe and Pb levels, relative to the first flush samples collected during regular-use before the pandemic.

## 2. Materials and Methods

### 2.1 Operation of a laboratory-scale premise water distribution system

A laboratory-scale premise water supply network with a 100 L overhead secondary storage was operated for 52 weeks through the monsoon, winter, and summer seasons, in that order. GI pipes (15 mm diameter), GI fixtures, and joints connected with threads, which are popular in the target region (MJP, 2012), were used to build this laboratory-scale premise water supply network. Throughout the experiment, the main supply line was in regular use, limiting water stagnation to the laboratory-scale water supply network. Water received from gravity-fed main supply lines connected to the OHT-based water distribution network was first stored in a 100 L secondary storage tank, chlorinated with bleaching powder (upto 2 ppm free chlorine concentration), and then supplied into the premise water supply network. Right after the secondary storage tank, the premise water supply network branched into two identical sections: 1D -flushed once daily, and 3D -flushed once in three days. The 1D and 3D further split into three lines, two of which (lines 1 and 3) were identical with a total length of 4.1 m and seven equidistant 90°elbows. The second pipeline (line 2) was 3.85 m long and had six equidistant 90°elbows. A tap at the end of the three lines was used for sampling and flushing the water (Figure S1). The flushing of fixtures (500 mL per tap at a flow rate of 4.7 GPM; calculated Reynold’s number >19,000) resulted in the withdrawal of 6% water by volume from the secondary storage tank for 1D and 12% when flushing under both 1D and 3D. The water was refilled into the secondary storage every 15-18 days when it was half empty, as is typical in the target region. The laboratory-scale premise water supply system was exposed to ambient temperature variations (5 − 39**°**C), as are the water supply systems in the area. The TOC in the water supplied into the secondary storage tank ranged from 4−7 mg/L (Table S6), which was comparable to the TOC of the water supplied during regular use before the pandemic (4−20 mg/L) (Bhaskar and Singh, 2021) (Table S7).

For microbiological analysis, 500 mL of the first flush was collected every twelve days from all the taps (both 1D and 3D) into autoclaved polypropylene bottles after flame-sterilizing the taps. The water samples were aseptically filtered through a 0.2 µm cellulose acetate filter paper (Axiva Sichem Biotech, India). The filter paper was aseptically folded using flame-sterilized forceps and stored at -20°C until further processing. At the end of its 52-week-long operation, the network was sacrificed to collect biofilm samples: each elbow (n= 7 for lines 1 and 3, n= 6 for line 2, Figure S2) and visibly corroded sections of the pipe surface (n= 24 for lines 1 and 3, and n = 21 for line 2, Figure S2) were swabbed with sterile cotton swabs for sampling the biofilms and stored in sterile screw-capped polypropylene tubes (Himedia Laboratories, Mumbai, India) at -20°C until further processing. Biofilms in taps (n=3 each for both 1D and 3D) were also sampled by swabbing a 2 cm section of the spout. The levels of the antibiotic resistance gene markers (*sul*1, *sul*2, *bla*OXA-1, *erm*F), Class 1 integron-integrase gene cassette marker (*int*I1) and DNA markers of opportunistic pathogens (*L. pneumophila* and *M. avium*) in these samples were analysed via qPCR. A recently reported year-long pre-pandemic investigation of the quality of the first flush of water from overhead tanks and buildings before the pandemic hereafter referred to as ‘OHT’ and ‘building’ samples in the targeted water distribution network (Bhaskar and Singh, 2021) served as a benchmark to compare the effect of 1D and 3D approaches on water quality, ARGs, and prevalence of opportunistic pathogens during the COVID19-related lockdown.

### 2.2 Water quality and heavy metal analysis

Physicochemical parameters of water, i.e. conductivity, pH, TOC and anions, were measured in the laboratory within 5-6 hours of sampling. Conductivity was measured using a WTW InoLab Cond 720® conductivity probe (Xylem Analytics, Oberbayern, Germany), and pH was measured by Toshcon pH meter® (Toshcon, Ajmer, India). Total organic carbon was measured on the Analytik Jena HT1300 TC/TN analyzer (Analytik Jena, Jena, Germany). Nitrate, sulfate, and chloride were measured using Metrohm ion chromatography (Metrohm, Herisau, Switzerland). For heavy metals analyses, nitric acid was added to the water samples to get pH <2 and stored at 4°C until analyzed for Cu, Pb, Fe, and Zn using ICP MS (ELAN DRC-e, Perkin Elmer, USA). The water quality index (WQI) (Tyagi et al., 2020) was calculated using the general water quality parameters (pH, conductivity, levels of chloride, nitrate, sulphate, Pb, Fe, Cu, and Zn).

### 2.3 DNA Extraction and molecular biological analysis

DNA was extracted using FastDNA Spin Kit for Soil® (MP Biomedicals, California, USA) according to the manufacturer’s instructions, with a minor modification at the cell-lysis step: TissueLyser (Qiagen, Hilden, Germany) was used at 25Hz for 30 seconds. The concentration of the DNA was determined through Qubit™ 4 Fluorometer (ThermoFisher Scientific, USA) following the manufacturer’s protocols. All water samples were diluted 1:10 with molecular biology grade – DNase-free water (Himedia Laboratories, Mumbai, India) to minimize quantitative polymerase chain reaction (qPCR) inhibition. Antibiotic resistance genes conferring resistance against sulphonamides (*sul*1 and *sul*2), beta-lactamase (*bla*OXA-1), and macrolides (*erm*F) were targeted as summarized in Table-S2 of SI using qPCR. All qPCR reactions were done in real-time PCR cycler, the Rotor-Gene Q® (Qiagen, Hilden, Germany) and the relevant details are available in Table S2. All primers and probes were synthesized by Eurofins Genomics (Bangalore, Karnataka, India). The standards for qPCR assays were prepared by inserting the target amplicon into pCR™ 2.1-TOPO® TA cloning vector using TOPO™ TA Cloning™ Kit for Sequencing (Invitrogen, California, USA) and transforming the vector into chemically competent *Escherichia coli* cells following the manufacturer’s protocol. The M13 amplicons in the successful clones were amplified and quantified using a NanoDrop™ One/One C Microvolume UV-vis spectrophotometer (ThermoFisher Scientific, Massachusetts, USA) after testing the specificity as described elsewhere (Bhaskar and Singh, 2021). The M13 amplicon was diluted serially over eight orders of magnitude (10^1^ – 10^8^ gene copies per µL), which were used as standards for the qPCR assays. Each qPCR run included a negative control in which molecular biology grade water was used instead of samples. Unless the negative controls gave no signal or its Ct value was higher than LOD, indicating that the signal in the negative control was below detection, the qPCR assay was discarded.

### 2.4 Statistical analysis

The ‘absolute level’ of the targeted gene marker was defined as the levels of its copy number in the sample as quantified by qPCR, while the ‘relative level’ of the relevant gene was defined as the ratio of absolute levels of the targeted genetic marker after normalizing it to the levels of 16S rRNA gene copies. The Shapiro-Wilk test was used to test the normality of data, and since the data was not normally distributed, only non-parametric tests were used throughout. One-tailed Wilcoxon signed-rank test was used to compare the differences in the levels of physicochemical parameters and log-transformed gene copy data. Augmented Dickey-Fuller test was done to test for the stationarity of the data over the course of the experiment. Statistical significance was set at p <0.05. R statistics® (version 4.0.3) (Core R Team, 2019) was used for statistical analysis, and the package ggplot2 (Hadley Wickham, 2009) was used for data visualization.

## 3. Results

### 3.1 Water quality changes under 1D and 3D approaches

The WQI was calculated following the CCME guidelines (Betis et al., 2020) using the parameters-pH, conductivity, and levels of chloride, nitrate, sulphate, Pb, Fe, Cu, and Zn. The higher the value of the WQI, the better the water quality. Overall, the WQI trended as building (WQI= 72) >1D (WQI= 58.4) and 3D (WQI= 58.3). Under both the 3D and 1D approaches, the water quality was poor (WQI= 45-59) relative to the water quality of (pre-pandemic) building samples, which was fair (WQI= 60-79). Heavy metals – Fe, Zn, and Cu - were significantly higher in the tap water in both 1D and 3D relative to building samples (p<0.05, One-tailed Wilcoxon signed-rank test) (Figures S3, S4, and S5). The Fe levels were higher in 1D relative to 3D (p=0.002, One-tailed Wilcoxon signed-rank test, Figure S3) and pre-pandemic building samples (p<0.05, One-tailed Wilcoxon signed-rank test, Figure S3). The Pb levels in tap water from both 1D and 3D were similar to that in the building samples (p>0.05, One-tailed Wilcoxon signed-rank test, Figure S6). The proportion of the samples that exceeded the WHO recommendations and BIS standards for drinking water (Bureau of Indian Standards, 2012; World Health Organization, 2017) for drinking water trended as for Pb (10 ppb) - 32% of 1D samples (30/93) > 31% of 3D samples (29/93) > building samples 29% (24/81) (Figure S6), and for Fe (300 ppb) - 48% (45/93) of 1D samples > 35% (33/93) of 3D samples > building samples 23% (19/81) (Figure S3).

While the feedwater was the same for both 1D and 3D, the conductivity of water was significantly higher in 3D (median = 370 µS/cm^2^) relative to 1D (median= 363 µS/cm^2^) (p= 0.02, One-tailed Wilcoxon signed-rank test, Figure S7). The TOC ranged from 2 − 16 ppm across all the tap water samples (n = 186) and trended as 3D (2 − 16 ppm) > 1D (1 − 13 ppm) > feedwater (4 − 7 ppm) (p= 0.03, One-tailed Wilcoxon signed-rank test, Figure S8). The levels of chloride (p=0.001, One-tailed Wilcoxon signed-rank test, Figure S9) and nitrate (p=0.02, One-tailed Wilcoxon signed-rank test; Figure S10) were higher under the 3D approach relative to 1D, while the sulphate levels were comparable under both 1D and 3D approaches (p>0.5, One-tailed Wilcoxon signed-rank test, Figure S11). The tap water was corrosive in 12% of 1D and 10% of 3D samples as indicated by the high levels (>0.5) of CSMR (Figure S12). In 1D, the CSMR of tap water correlated with Zn (r= 0.29) (Figure 5A, Table S3), and TOC correlated with Fe (r= 0.27), Pb (r= 0.39), Cu (r= 0.41), and Zn (r= 0.25). In the tap water under the 3D approach, the CSMR correlated with Zn (r= 0.21); and TOC levels correlated with Fe (r= 0.22), Pb (r= 0.44), Cu (r= 0.38), and Zn (r= 0.42); conductivity correlated with Fe (r= −0.21), Pb (r= −0.26), Cu (r= −0.45), and Zn (r= −0.21) (Figure 5B, Table S4). The pipe sections under the 1D and 3D approaches were visibly corroded (Figure S3).

### 3.2 Trends in the 16S rRNA gene copy number under 1D and 3D approaches

Since the 16S rRNA gene copy numbers obtained via qPCR account for both live and dead microbes with intact DNA, the levels of 16S rRNA gene copy numbers were employed only to track overall trends in the total bacterial count. The levels of 16S rRNA gene copies in both the tap water and the biofilms samples under the 3D approach were 2-3 orders of magnitude higher relative to the 1D approach, the OHT, and the building samples (p<0.05, One-tailed Wilcoxon signed-rank test, Figure 1 & S13). There was no difference in the levels of 16S rRNA gene copies in the tap water from 1D and the buildings under routine occupation before the pandemic (p= 0.4, One-tailed Wilcoxon signed-rank test). The levels of 16S rRNA gene copies in the tap water varied throughout the duration of the operation of the laboratory-scale premise water supply network (p=0.01, Augmented Dickey-Fuller test) under both the 1D and 3D approaches: During the first 120 days of operation, which coincided with the monsoon month (water temperature: 23°C – 25°C; July to September), the levels of 16S rRNA gene copies increased by an order of magnitude in 1D, and 3-4 orders of magnitude in 3D (Figure 1A). Between day 120 - day 230 of the operation, which coincided with the winter months (water temperature: 12°C – 16°C, late October to March), the levels of 16S rRNA gene copies decreased an order of magnitude in 1D (Figure 1A, p= 0.001, One-tailed Wilcoxon signed-rank test), and had a slight but statistically insignificant decrease in 3D (Figure 1A, p= 0.08, One-tailed Wilcoxon signed-rank test). Over the next 136 days (day 230 – day 365 of the operation), which coincided with summer months (water temperature: 28°C – 35°C; March to June), the levels of 16S rRNA gene copies increased two orders of magnitude in both 1D (p= 0.001, One-tailed Wilcoxon signed-rank test) and 3D (p= 0.002, One-tailed Wilcoxon signed-rank test) (Figure 1A). The levels of 16S rRNA gene copies in the tap water weakly but significantly correlated with the levels of Fe in 1D (r= 0.25, Spearman’s rank correlation); and with chloride (r=0.29) and sulphate (r=0.29) in 3D (Figure 5A).

**Fig. 1.**
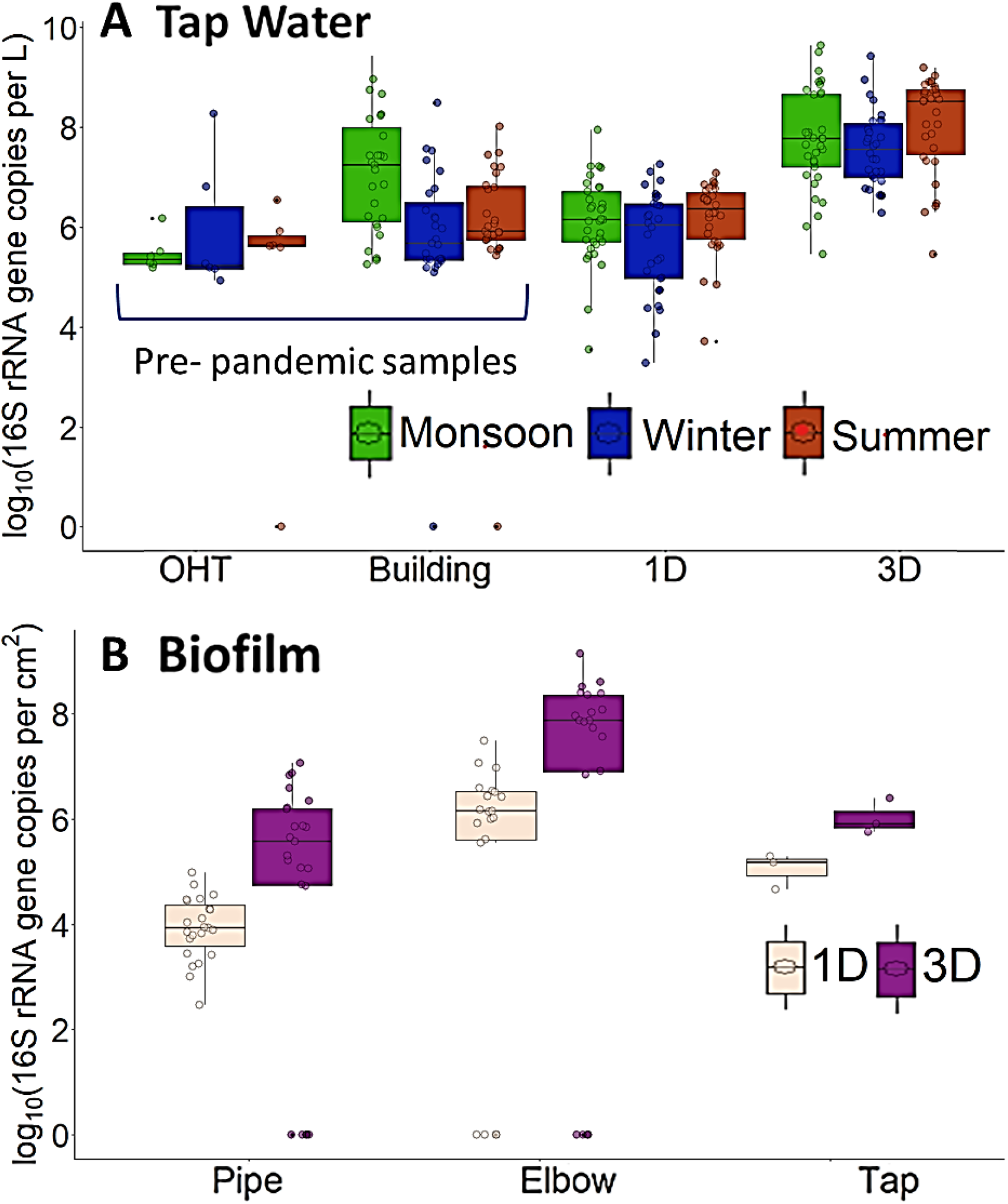
The levels of 16S rRNA gene copies in tap water samples collected from **A**. pre-pandemic OHT, buildings, and laboratory scale premise water supply network in three seasons (summer, monsoon and winter) and, **B**. Biofilm samples swabbed from surfaces of pipes, elbows and taps subjected to 1D and 3D approaches; Prefixes 1D and 3D refer to water drawn once per day and once in three days respectively

The levels of 16S rRNA gene copies per unit area in the biofilms from 3D were 2-3 orders of magnitude higher relative to 1D (p<0.05, One-tailed Wilcoxon signed-rank test, Figures 1B & S13B). Within the laboratory-scale premise water distribution system, the biofilm samples from the elbows had two orders of magnitude higher levels of 16S rRNA gene copies relative to visibly-corroded pipe-sections for both 1D (p<0.05, One-tailed Wilcoxon signed-rank test, Figure 1B) and 3D approaches (p=0.0003, One-tailed Wilcoxon signed-rank test, Figures 1B & S13B). The biofilm sample taken from the elbows that were farthest from the secondary storage tank under the 3D approach, and elbows that were second to last from the secondary storage tank under the 1D approach had the highest level of 16S rRNA gene copies per unit area (Figures S14A & S15A).

### 3.3 Levels of antibiotic resistance gene markers

#### 3.3.1 Tap water

Of the ARGs targeted in this study - *sul*1, *sul*2, *erm*F, *bla*OXA-1 - only *sul*1 and *int*I1 were quantifiable (SI). Overall, the absolute levels of *sul*1 in the tap water under 3D (0 − 10^7.68^ genes copies/L) were 1-2 orders of magnitude higher relative to 1D (0 – 10^6.74^ gene copies/L) (p<0.05, One-tailed Wilcoxon signed-rank test, Figure S16A), and two orders of magnitude higher relative to building samples (10^3.4^ – 10^6.5^ gene copies/L) (p<0.05, One-tailed Wilcoxon signed-rank test, Figure S16A). While the *sul*1 levels in tap water from the 1D approach were comparable to the pre-pandemic levels in building samples, during winter, the *sul*1 levels in 1D were significantly higher relative to the pre-pandemic building samples (p<0.01, One-tailed Wilcoxon signed-rank test, Figure 2A). When normalized with 16S rRNA gene copy number, the relative levels of *sul*1 were significantly higher in 1D relative to 3D (p<0.05, One-tailed Wilcoxon signed-rank test, Figure S17A), with the highest levels recorded in winter, when they were nearly an order of magnitude higher relative to 3D (Figure S17A). The absolute levels of *int*I1 in 1D (0 – 10^5.9^ gene copies/L) and 3D (0 – 10^6.8^ gene copies/L) approaches were comparable relative to the building samples (0 – 10^5.8^ gene copies/L) (p>0.05, One-tailed Wilcoxon signed-rank test, Figure S18A). The relative levels of *int*I1 were significantly higher under the 1D approach (−6.2 to −0.15 on the log10 scale) relative to the 3D approach (− 7.9 to −0.76 on log10 scale), trending similarly as the relative levels of *sul*1. The relative levels of *int*I1 in winter and summer seasons in 1D (− 6.2 to −2.8 on the log10 scale) were 2-3 orders of magnitude higher compared to the building samples (− 6.8 to −0.79 on the log10 scale) (p<0.05, One-tailed Wilcoxon signed-rank test, Figure 3A). During the monsoon season, the absolute and relative levels of *int*I1 under the 1D approach and in building samples were comparable (Figure 3A).

**Fig. 2.**
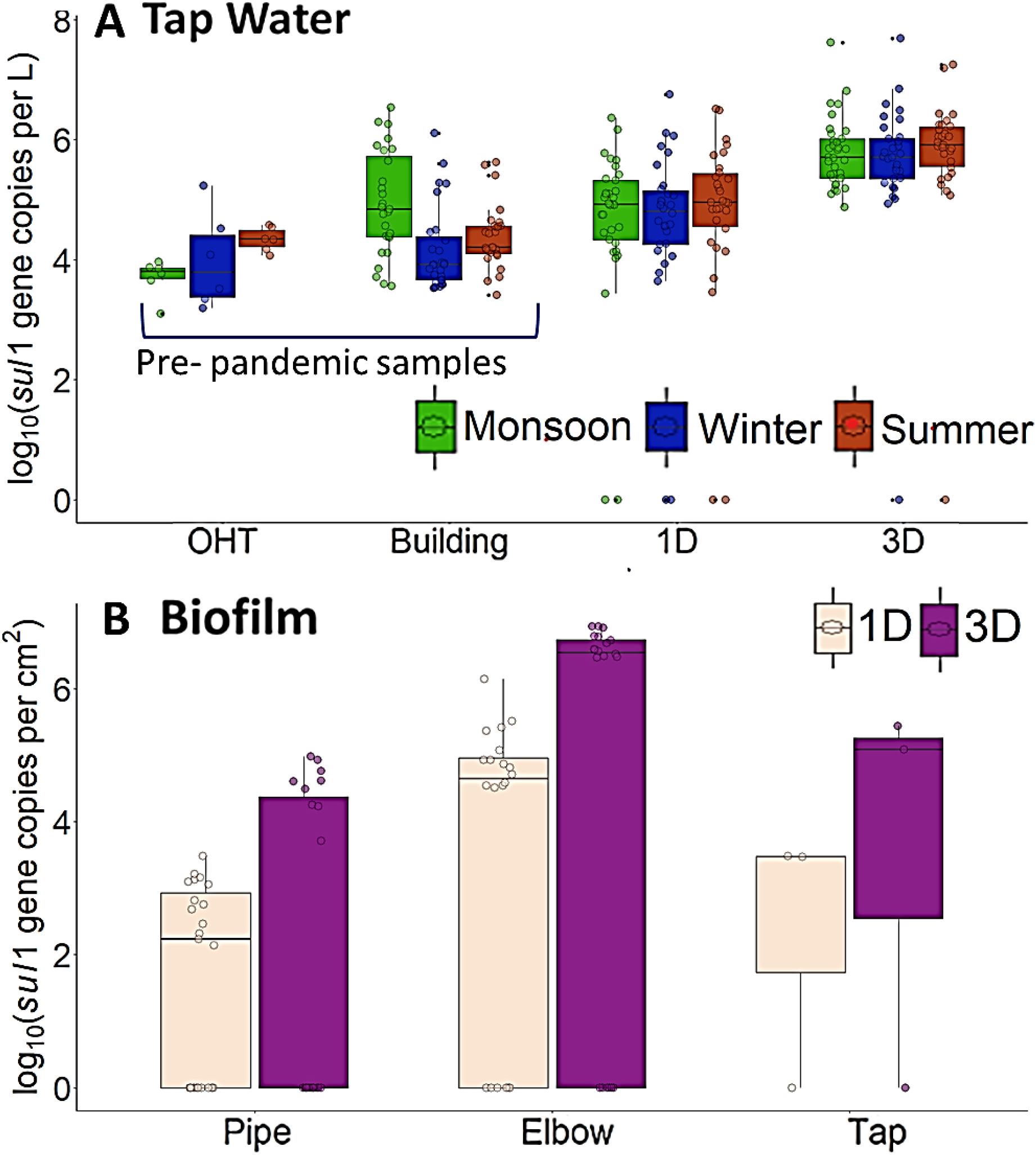
The levels of *sul*1 in **A**. tap water samples collected from pre-pandemic OHT, buildings and laboratory scale premise water supply network in three seasons (summer, monsoon and winter) and, **B**. biofilm samples swabbed from surfaces of pipes, elbows and taps subjected to 1D and 3D approaches; Prefixes 1D and 3D refer to water drawn once per day and once in three days respectively

**Fig. 3.**
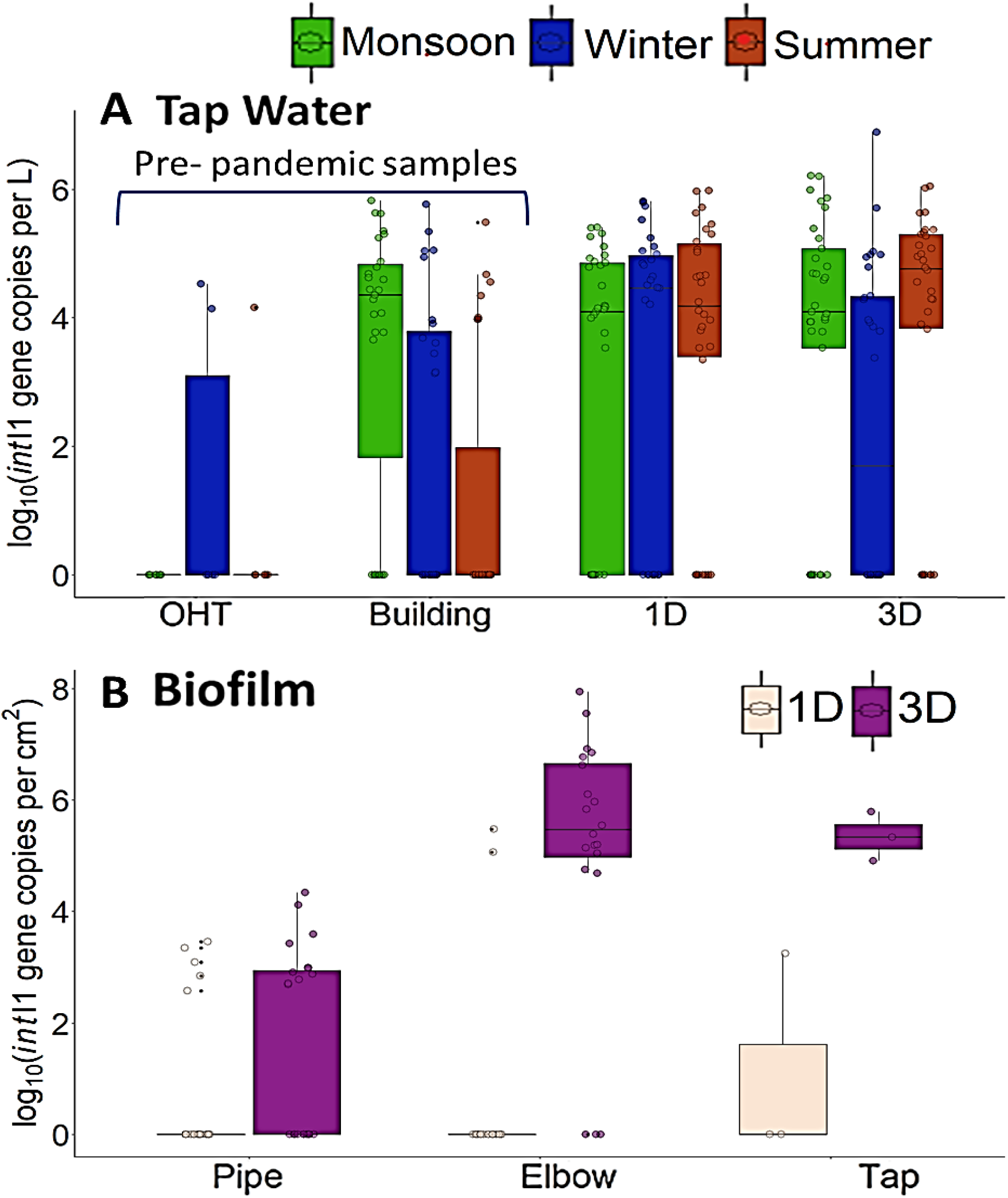
The levels of *int*I1 gene copies in water samples collected from **A**. pre-pandemic OHT, buildings and laboratory scale premise water supply network in three seasons (summer, monsoon and winter) and, **B**. biofilm samples swabbed from surfaces of pipes, elbows and taps subjected to 1D and 3D approaches; Prefixes 1D and 3D refer to water drawn once per day and once in three days respectively

Similar to the 16S rRNA gene copy levels, the absolute levels of *sul*1 and *int*I1 varied throughout the year-long operation. In both 1D, the absolute levels of both *sul*1 and *int*I1 increased slightly for the first 230 days, followed by a slight dip during the winter season and an insignificant rise during the summer season (p>0.5, One-tailed Wilcoxon signed-rank test), while in 3D a significant drop was seen in winter for *int*I1 (p<0.05, One-tailed Wilcoxon signed-rank test) (Figure 2A & 3A). While the absolute (0 – 10^7.68^ gene copies/L) and relative (−6.8 to 0.3 on log10 scale) levels of *sul*1 in 3D were consistently higher relative to 1D throughout the year-long operation of the laboratory-scale premise water supply network, the absolute (0 – 10^6.5^ gene copies/L) and relative (− 6.2 to 0.6 on log10 scale) levels of *int*I1 in 1D and 3D were different only during the last 135 days of the operation when the levels of *int*I1 in 1D slightly exceeded the levels in 3D (Figure S19A). The relative levels of *int*I1 increased over the year in 1D and decreased somewhat after the winter season in 3D (after 210 days) (Figure S19A).

Under the 1D approach, the relative levels of *sul*1 correlated negatively with TOC levels (r= −0.22) (Figure 5A, Table S3), while the relative levels of *int*I1 correlated with chloride (r= 0.30) and nitrate (r= 0.28) levels, and negatively with pH (r= −0.2) (Figure 5A, Table S3). In 3D, the relative levels of *sul*1 and *int*I1 in tap water did not correlate with any of the measured parameters (Figure 5B, Table S4).

#### 3.3.2 Biofilms

The absolute levels of both *sul*1 and *int*I1 in the biofilms were higher in 3D relative to 1D: an order of magnitude higher for *sul*1 and 3 orders of magnitude higher for *int*I1 (p<0.05, One-tailed Wilcoxon signed-rank test, Figures S16B and S18B). Within the premise water supply network, the absolute levels of *sul*1 trended as elbows (0− 10^6.9^ gene copies/cm^2^) > taps (0− 10^5.4^ gene copies/cm^2^) > visibly corroded pipe-sections (0− 10^4.9^ gene copies/cm^2^) in both 1D and 3D (p≤0.05, One-tailed Wilcoxon signed-rank test, Figure 2B). The absolute levels of *int*I1 in biofilms samples from 3D trended as elbows (0− 10^7.9^ gene copies/cm^2^) > taps (0− 10^5.7^ gene copies/cm^2^) > visibly corroded pipe sections (0− 10^4.3^ gene copies/cm^2^) (p<0.05, One-tailed Wilcoxon signed-rank test, Figure 3B). Similarly, in 1D, the absolute levels of *int*I1 in biofilms trended as elbows (0− 10^5.4^ gene copies/cm^2^) > visibly corroded pipe-sections (0− 10^3.4^ gene copies/cm^2^) > taps (0− 10^3.2^ gene copies/cm^2^) (p<0.05, One-tailed Wilcoxon signed-rank test, Figure 3B). The absolute *sul*1 and *int*I1 levels in elbows and pipe sections did not vary with the distance from the secondary storage tank under 1D and 3D approaches (Figures S13B, S13C, and S14B, S14C). The relative levels of *sul*1 in 1D were comparable in samples taken from visibly corroded pipe sections (−4 to 0.4 on log10 scale), elbows (−6 to 0.1 on log10 scale) and taps (−4 to −0.5 on log10 scale) at the user-end (Figure S17B), while in 3D, it trended as elbows (−7 to 0.5 on log10 scale) > visibly corroded pipe-sections (− 6 to 0.3 on log10 scale)> taps (−5 to 0.1 on log10 scale) (Figure S17B). The relative levels of *int*I1 in the biofilms in 1D trended as elbows (−6.8 to −0.2 on log10 scale) < taps (−4 to −1.3 on log10 scale) and pipe-sections (−4 to −0.01 on log10 scale) (Figure S19B), and in 3D, the absolute levels of *int*I1: taps. (−0.8 to 0.04 on log10 scale) >elbows (−7.4 to 0.04 on log10 scale) >pipe-sections (−6.4 to −1.1 on log10 scale) (Figure S19B).

### 3.4 Pathogens in the simulated water distribution system

The tap water and biofilm samples were analyzed for the DNA markers of *L. pneumophila, M. avium*, and *E. coli*, amongst which only the DNA marker for *L. pneumophila, mip*, was detected. When detected, the *mip* gene levels in tap water were not statistically different in 1D, 3D, and building samples (p>0.05, One-tailed Wilcoxon signed-rank test, Figure S20A). However, the prevalence of *mip* in tap water samples trended as 1D (77/93) > 3D (70/93) > building samples (50/81). The *mip* levels in tap water under the 1D approach correlated with Fe (r= 0.30), Pb (r= 0.33), Cu (r= 0.27), and Zn (r= 0.32) (Figure 5A, Table S3). Whereas, under the 3D approach, the *mip* levels correlated with the levels of Fe (r=0.28), Pb (r= 0.23), Zn (r= 0.27), and sulphate (r= −0.30) (Figure 5B, Table S4). The *mip* levels changed significantly throughout the year in both 3D and 1D, similar to the variations observed in the building samples (Bhaskar and Singh, 2021) (Figure 4A). During the winter season, both the 1D (24/93) and 3D (20/93) approaches had a much higher prevalence of *mip* relative to the building samples 6% (2/31) (Figure 4A). During summer and monsoon seasons, the prevalence of *mip* was in 1D, and 3D was comparable to the building samples. The levels of *mip* in the biofilm samples were similar in both 1D (0-10^5.55^ gene copies/cm^2^) and 3D (0-10^5.5^ gene copies/cm^2^) (p<0.05, One-tailed Wilcoxon signed-rank test, Figure S20B), and trended as elbows (0-10^5.5^ gene copies/cm^2^) > pipe-sections (0-10^5.3^ gene copies/cm^2^) > taps (0-10^3.9^ gene copies/cm^2^) (p<0.05, One-tailed Wilcoxon signed-rank test) (Figure 4B).

**Fig. 4.**
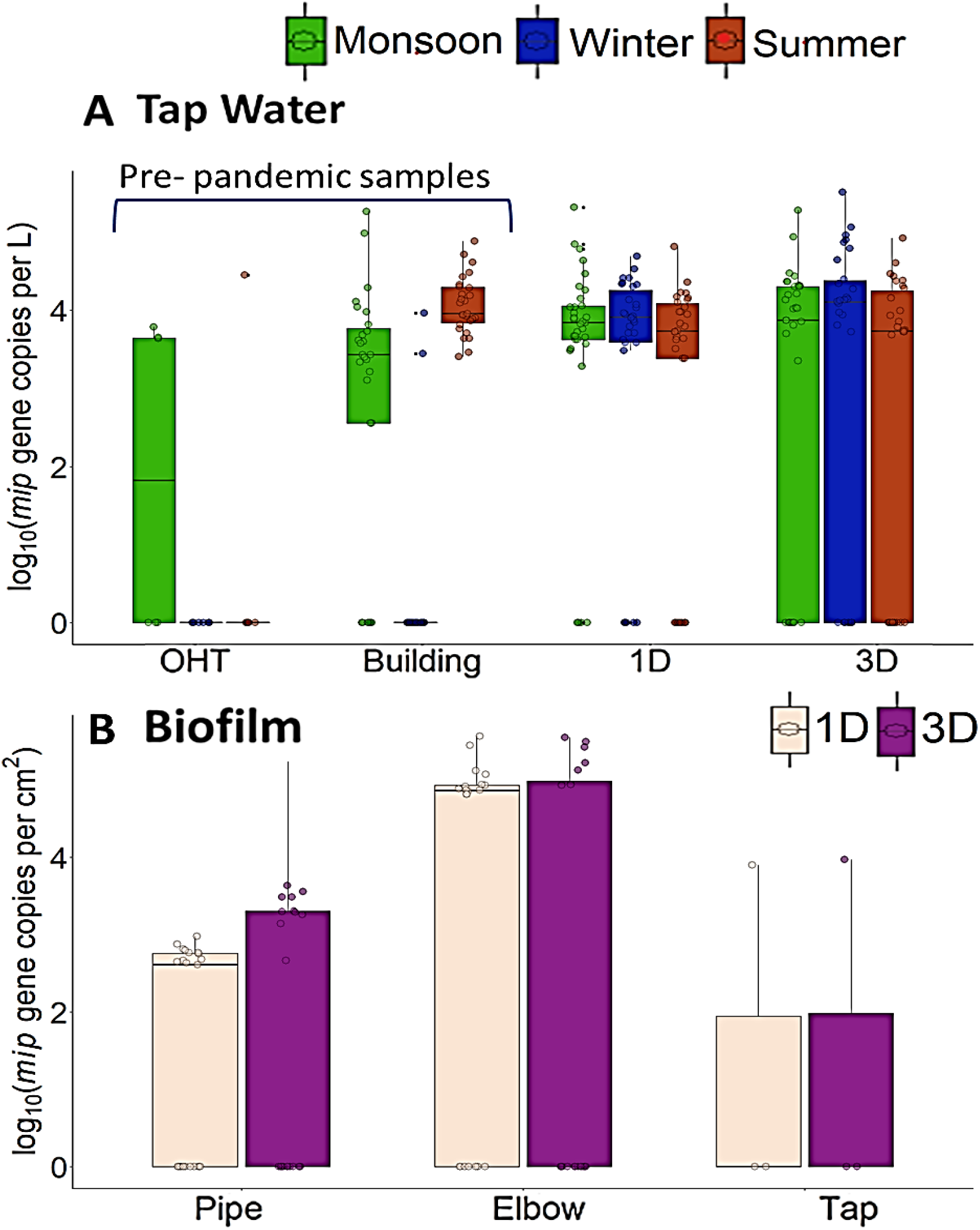
The levels of *mip* gene copies in tap water samples collected from **A**. pre-pandemic OHT, buildings and laboratory scale premise water supply network in three seasons (summer, monsoon and winter) and, **B**. biofilm samples swabbed from surfaces of pipes, elbows, and taps subjected to 1D and 3D approaches; Prefixes 1D and 3D refer to water drawn once per day and once in three days respectively

In 1D biofilms, the absolute *mip* levels correlated with absolute *sul*1 levels (r= 0.30), whereas in the tap water, the levels of *mip* gene under the 1D approach negatively correlated with absolute levels of *sul*1 (r= −0.29). Only in 1D elbows, the absolute *mip* correlated with the distance of the elbow from the secondary storage tank (r= 0.61) (Figure 5A).

**Fig. 5.**
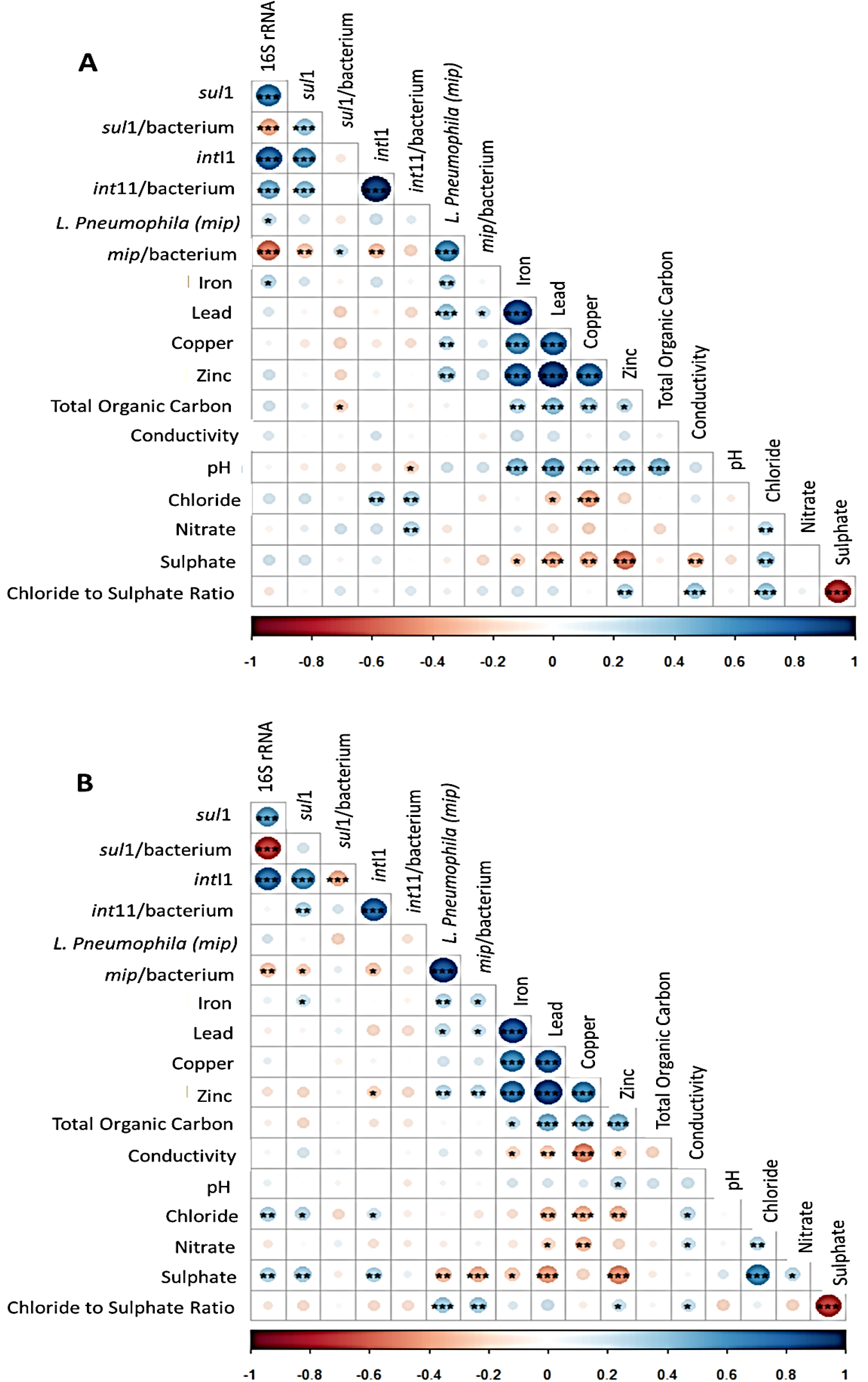
Correlogram of Spearman’s rank correlation coefficients computed between the relative concentration of targeted genes and physicochemical parameters in all samples in **A**. 1D and, **B** 3D laboratory scale premise water supply network (*: P-value < 0.05, **: P-value < 0.01, ***: P-value < 0.001)

## 4. Discussion

### 4.1 Once a day flushing of taps (1D) was sufficient to contain any increase in the total bacterial count in water, but the water quality still worsened

The water quality index reduced from ‘fair’ in pre-pandemic building samples to ‘poor’ in 1D and 3D (Betis et al., 2020). Water from taps flushed once a day (1D) had similar levels of 16S rRNA gene copies over the year as the tap water building samples collected under routine occupation before the pandemic. Short-term studies which recommend flushing the water regularly to prevent bacterial growth have had similar observations (Hozalski et al., 2020; Rhoads and Hammes, 2021). In this year-long experiment, it took flushing 6% of water stored in the secondary storage tank every day (1D) to keep the 16S rRNA gene copy levels in the first flush comparable to the first flush from the buildings under regular use a year before the pandemic. However, when the frequency of flushing the taps was decreased to once in three days (3D), the total bacterial count increased substantially both in the tap water and biofilms inside the laboratory-scale premise water distribution system (2-3 orders of magnitude). Thus, the frequency at which the water is flushed is critical in containing the bacterial count in the tap water and biofilms. Flexibility in the frequency of preventative flushing is especially relevant for situations where it would be more feasible or convenient to flush the water less frequently in case of similar lockdowns in future due to weaker follow-up of regulations or lack of resources or inability to ensure the safety of the person responsible for flushing the taps for the duration of long-term building shutdown (Hozalski et al., 2020).

Both 1D and 3D saw an increase in the levels of total bacterial count during the monsoon season – similar to the pre-pandemic building samples. However, this could also be attributed to the initial bacterial colonization of the laboratory-scale premise water supply system (Douterelo et al., 2016a) and possibly the seasonal increase in feedwater as observed during the monsoon season in the pre-pandemic building samples (Bhaskar and Singh, 2021). Unlike the 3D approach, the 1D approach saw a dip of an order of magnitude in winter in 16S rRNA gene copy levels, similar to the pre-pandemic building samples that saw a substantial reduction in the 16S rRNA gene copies levels (Bhaskar and Singh, 2021). The changes in ambient temperature still impacted any additional bacterial growth in 1D due to stagnation and secondary storage was still impacted by similar to the pre-pandemic building samples.

While the 1D approach managed to contain the increase in the levels of 16S rRNA gene copies in the first flush, the water still had elevated levels of heavy metals, which did not meet the water quality standards. Since the levels of heavy metals - Fe, Pb, Zn and Cu - in the tap water correlated with the TOC and CSMR of the water supplied to the secondary storage tank, it is possible that the high TOC and CSMR of water-fueled corrosion and subsequently the increase in the levels of heavy metals. Further, the levels of 16S rRNA gene copies in water correlated significantly with Fe in the water, suggesting corrosion-fueled microbial growth.

The first flush was collected and analyzed as both the mandate and the practice of removing the first flush are absent in the target region (Bhaskar and Singh, 2021). Infact, upon resumption of regular use after the lifting of pandemic-related lockdowns in the (academic) buildings in the target region, the water was not flushed before the resumption of occupation, as advised by Richard et al., 2020.

### 4.2 The anthropogenic antibiotic resistance gene marker levels increased in both tap water and biofilms in both 1D and 3D

In both fully pressurized and gravity-fed systems, corrosive water has been linked with opportunistic pathogens and increased bacterial count, namely an increase in anthropogenic antibiotic resistance gene markers (Bhaskar and Singh, 2021). The DNA markers of sulfonamide resistance gene, *sul*1, and class 1 integron-integrase gene, *int*I1, quantified in this study are referred to as the anthropogenic antibiotic resistance gene markers due to strong evidence of their association with the human-impact driven increase in antibiotic resistance levels in the environment (Davis et al., 2020). Class 1 integron-integrase cassettes are not only linked with the horizontal gene transfer of antibiotic resistance (Gillings et al., 2015) but also with the horizontal transfer of other genes, such as metal resistance genes, that might give bacteria a competitive advantage, which is relevant in case of corrosive water with elevated heavy metals levels. A previous study on *sul*1 and *int*I1 in the tap water supplied by an overhead-tank-based water network – that also supplied water to the laboratory-scale premise water supply network – observed an increase in relative levels of *int*I1 but not of *sul*1 gene copies in the water distribution network (Bhaskar and Singh, 2021). Under both the 1D and 3D approaches, the absolute and relative levels of the anthropogenic antibiotic resistance gene markers in tap water increased relative to the pre-pandemic building samples during winter and summer. Since the experiment commenced at the onset of the monsoon, the comparable levels of anthropogenic antibiotic resistance gene markers in 1D and the building samples during the monsoon could be attributed to initial colonisation in 1D and higher contamination in building samples. Overall, the relative levels of *sul*1 and *int*I1 in 3D were lower than in 1D. In 1D, relatively more regular replacement of nutrients and residual disinfectant (a co-selective agent) and higher heavy metal levels (potential co-selective agent) - could have reduced the advantage of forgoing *sul*1.

As the relative levels of *int*I1 increased in 1D over time, the efficacy of flushing water daily to protect the potability of water could vary with the duration of the lockdown. Thus, it is crucial to monitor the effects of flushing water regularly for long-term building-specific shutdowns. On the other hand, in the relatively nutrient-rich and stagnant environment of biofilms, which are conducive to bio-corrosion (Seiler and Berendonk, 2012), the relative levels of *int*I1 were higher under the 3D approach than under the 1D approach.

The CSMR of the tap water correlated with the relative levels of *sul*1 and *int*I1, indicating that the corrosivity of water (Kimbell et al., 2020) is linked with the increase of the anthropogenic antibiotic resistance gene markers. Elevated CSMR has been linked to galvanic corrosion and subsequent release of Fe and Pb (Kogo et al., 2017), which are co-selective agents (Kogo et al., 2017). Since the relative levels of *sul*1 and *int*I1 in biofilms correlated with each other, it is possible that the increase in the relative levels of *sul*1 levels in the biofilms was linked with class 1 integron-integrase mediated horizontal gene transfer within the nutrient-rich biofilms (Kogo et al., 2017) or was driven by confounding factors.

### 4.3 The DNA marker of opportunistic pathogens was more commonly detected despite flushing

The DNA marker for *L. pneumophila, mip*, was detected in both the tap water and biofilms of both 1D and 3D (10^3^ to 10^5^ gene copies/L), and it is comparable to the levels reported elsewhere (Hamilton et al., 2019). Since the levels of *L. pneumophila* plateau at the infectious point (Hamilton et al., 2019), it is not surprising that the levels of *mip* were comparable in 1D, 3D, and building samples, whenever *mip* was detected. Risk analysis elsewhere (Hamilton et al., 2019) suggested that a higher number of exposure events in the case of *L. pneumophila* can substantially increase the risk of infection. Hence, once the *L. pneumophila* levels reach the infectious point, the number of times it is detected in water, which would directly affect the exposure, becomes important from a risk perspective.

The presence of the DNA marker of *L. pneumophila* in tap water alone does not indicate the presence of the live and viable pathogen. However, the trends in the prevalence of its DNA marker in tap water suggest trends in its growth or more frequent release of the DNA marker into tap water. Under the 1D and 3D approaches, the levels of the DNA marker of *L. pneumophila* were higher than in pre-pandemic building and OHT samples, indicating failure of flushing to prevent increased colonization by *L. pneumophila*. Within the biofilms, there was no significant difference in the prevalence and levels of *mip* in 1D vs. 3D. Our observations reveal the inefficacy of the recommendation of flushing frequently to control the growth of opportunistic pathogens in low-use or low-flow outlets (European Working Group for Legionella Infections, 2017). Even after replacing the water, the pathogens in the biofilms could continue to proliferate and eventually release into the water (Hozalski et al., 2020). It is important to note that the levels of *L. pneumophila* in first flush can be comparable to the levels in tap water after complete flushing (Wang et al., 2012).

Fe is a crucial cofactor for the growth of *L. pneumophila* (M. Christopher, 2016) and can also serve as either an electron donor or acceptor (Appenzeller et al., 2005) for the bacterial community in drinking water supply network. Interestingly, the levels of Fe in tap water correlated significantly with the levels of 16S rRNA gene copies and the DNA marker of *L. pneumophila* in both 1D and 3D. Major outbreaks of *L. pneumophila* have been associated with corrosive tap water elsewhere (Martin et al., 2020; Rhoads et al., 2020). Chlorination of water in storage tanks was alone entrusted with ensuring potable water at the user end in both the targeted region and the study presented herein. Daily flushing was insufficient to contain the increase in the prevalence of the DNA marker of *L. Pneumophila*. Our study also demonstrated that adequate chlorination in the overhead and secondary storage tank was insufficient to ensure safe water in taps despite the 1D approach.

### 4.4 Bacterial presence in the laboratory-scale premise-water supply system was highest in the elbows

Within the laboratory-scale premise water supply network, the biofilms in the elbows had the highest levels of 16S rRNA gene copies per unit area, anthropogenic antibiotic resistance gene markers - *sul*1 and *int*I1, and the DNA marker for *L. pneumophila* relative to the biofilms in the taps and the pipe-sections that were visibly corroded. The elbow bends are responsible for head loss and face extremes in fluid velocities (Muftah, 2014). Typically, the inner bend of the elbow will face a lower local water velocity in contrast to the outer bend. Given the drastic changes in water velocities at the elbows, they are also more susceptible to corrosion and related wear and tear. That the elbows had higher levels of microbial activity than sections of pipes further underlines that water flow patterns within the distribution system impact the water quality within the premises that utilize secondary storage and are connected to the OHT-based drinking water supply network. It was also only in elbows and taps where the relative levels of *int*I1 in the biofilms were highest. Since the elbows are more susceptible to corrosion, the increase in the relative levels of *int*I1 in elbows could be driven by corrosion (Baker-Austin et al., 2006; Pal et al., 2015).

In the biofilms, the absolute levels of *mip* correlated with absolute levels of *sul*1 which, while cannot suggest antibiotic resistance in *L. pneumophila*, shows similar trends in the growth of opportunistic pathogen and the levels of *sul*1. In tap water, however, the levels of *mip* correlated negatively with the absolute levels of *sul*1, indicating that antibiotic resistance may deselect in the low-nutrient tap water, as seen elsewhere (Bhaskar and Singh, 2021); the same cannot be said for opportunistic pathogens. The *mip* levels in biofilms positively correlated with the distance between the pipe-section and elbows from the secondary storage tank under both 1D and 3D approaches, suggesting a link between pressure head loss and growth of the opportunistic pathogen. The relatively small size of the laboratory-scale premise water supply network could result in an underestimation of the actual impact of 1D and 3D approaches that are to be expected in premise water distribution systems in the context of opportunistic pathogens. On the other hand, in the context of anthropogenic antibiotic resistance gene markers, the relative and absolute levels of *sul*1 and *int*I1 did not change with the distance of biofilms from the secondary storage tanks.

## 5. Conclusions

The prospect of using regular flushing to prevent bacterial growth, increased exposure to opportunistic pathogens, and increased overall antibiotic resistance in biofilms and tap water was investigated for premises with secondary storage that receive water from gravity-fed drinking water distribution system, as is the case in many developing countries. While flushing 6% of the water once a day from the taps prevented the increase in the levels of 16S rRNA gene copies, the water quality deteriorated in terms of heavy metals, anthropogenic antibiotic resistance gene markers, and prevalence of DNA marker of *L. pneumophila*. Overall, more extended stagnation in 3D led to an increase in absolute levels of the *sul*1, *int*I1, and 16S rRNA gene copy numbers, and the relative levels of *int*I1. Chlorinating water in the storage tanks was insufficient to prevent microbial growth and water quality deterioration. While the water can be flushed out of the distribution systems and storage tanks, prior to re-occupation of the premises, the antibiotic resistance gene markers and possibly opportunistic pathogens in biofilms and corrosion-related damage to the infrastructure due to stagnation could threaten public health.

## Data Availability

All data produced in the present work are contained in the manuscript

## Data Availability

All data produced in the present work are contained in the manuscript

## Acknowledgments

The authors would like to thank Jahnavi Kurasam, Sourabh Dixit, and Sonali Gupta for help during sampling, flushing and filling the storage tank; and Navneet Kumar Saini and Jitendra Singh Meena for sharing information related to the OHT-based DWDS investigated in this study. The authors declare no competing interest.

## Appendix

**Supplementary Materials**

Supplementary material associated with this article can be found online.

## References

Appenzeller, B.M.R., Yan, C., Jorand, F., Block, J., 2005. Advantage Provided by Iron for Escherichia coli Growth and Cultivability in Drinking Water 71, 5621–5623. https://doi.org/10.1128/AEM.71.9.5621

Baker-Austin, C., Wright, M.S., Stepanauskas, R., McArthur, J. V., 2006. Co-selection of an-tibiotic and metal resistance. Trends Microbiol. 14, 176–182. https://doi.org/10.1016/j.tim.2006.02.006

Betis, H., St-Hilaire, A., Fortin, C., Duchesne, S., 2020. Development of a water quality in-dex for watercourses downstream of harvested peatlands. Water Qual. Res. J. 55, 119–131. https://doi.org/10.2166/wqrj.2020.007

Bhaskar, D., Singh, G., 2021. Increase in anthropogenic antibiotic resistance markers in water supplied by an overhead tank based-water distribution system. Environ. Sci. Water Res. Technol. https://doi.org/10.1039/d1ew00267h

Bureau of Indian Standards, 2012. Indian Standard: Drinking water-Specification (second revision).

Cookson, M.D., Stirk, P.M.R., 2019. Impact of Tank Material on Water Quality in Household Water Storage Systems in Cochabamba, Bolivia.

Core R Team, 2019. A Language and Environment for Statistical Computing. R Found. Stat. Comput. 2, https://www.R--project.org.

Davis, B.C., Riquelme, M.V., Ramirez-Toro, G., Bandaragoda, C., Garner, E., Rhoads, W.J., Vikesland, P., Pruden, A., 2020. Demonstrating an Integrated Antibiotic Resistance Gene Surveillance Approach in Puerto Rican Watersheds Post-Hurricane Maria. Envi-ron. Sci. Technol. 54, 15108–15119. https://doi.org/10.1021/acs.est.0c05567

Douterelo, I., Husband, S., Loza, V., Boxall, J., 2016a. Dynamics of biofilm regrowth in drinking water distribution systems. Appl. Environ. Microbiol. 82, 4155–4168. https://doi.org/10.1128/AEM.00109-16

Douterelo, I., Jackson, M., Solomon, C., Boxall, J., 2016b. Microbial analysis of in situ bio-film formation in drinking water distribution systems: implications for monitoring and control of drinking water quality. Appl. Microbiol. Biotechnol. 100, 3301–3311. https://doi.org/10.1007/s00253-015-7155-3

EPA, 2021. Information on Maintaining or Restoring Water Quality in Buildings with Low or No Use.

Gillings, M.R., Gaze, W.H., Pruden, A., Smalla, K., Tiedje, J.M., Zhu, Y.G., 2015. Using the class 1 integron-integrase gene as a proxy for anthropogenic pollution. ISME J. 9, 1269–1279. https://doi.org/10.1038/ismej.2014.226

Hadley Wickham, 2009. ggplot2: Elegant Graphics for Data Analysis. Springer-Verlag New York. https://doi.org/10.1007/978-0-387-98141-3

Hamilton, K.A., Hamilton, M.T., Johnson, W., Jjemba, P., Bukhari, Z., Lechevallier, M., Haas, C.N., Gurian, P.L., 2019. Risk-Based Critical Concentrations of Legionella pneu-mophila for Indoor Residential Water Uses. https://doi.org/10.1021/acs.est.8b03000

Hozalski, R.M., Lapara, T.M., Zhao, X., Kim, T., Waak, M.B., Burch, T., Mccarty, M., 2020. Flushing of Stagnant Premise Water Systems after the COVID-19 Shutdown Can Re-duce Infection Risk by Legionella and Mycobacterium spp. Environ. Sci. Technol. 54, 15914–15924. https://doi.org/10.1021/acs.est.0c06357

Kimbell, L.K., Wang, Y., McNamara, P.J., 2020. The impact of metal pipe materials, corro-sion products, and corrosion inhibitors on antibiotic resistance in drinking water distri-bution systems. Appl. Microbiol. Biotechnol. 104, 7673–7688. https://doi.org/10.1007/s00253-020-10777-8

Kogo, A., Payne, S.J., Andrews, R.C., 2017. Comparison of three corrosion inhibitors in sim-ulated partial lead service line replacements. J. Hazard. Mater. 329, 211–221. https://doi.org/10.1016/j.jhazmat.2017.01.039

Li, M., Liu, Z., Chen, Y., Hai, Y., 2016. Characteristics of iron corrosion scales and water quality variations in drinking water distribution systems of different pipe materials. Wa-ter Res. 106, 592–603. https://doi.org/10.1016/j.watres.2016.10.044

M. Christopher, A.M.L.S., 2016. An update on iron acquisition by Legionella pneumophila: new pathways for siderophore uptake and ferric iron reduction. Physiol. Behav. 176, 100–106. https://doi.org/10.2217/fmb.15.21.An

Martin, R.L., Strom, O.R., Pruden, A., Edwards, M.A., 2020. Interactive effects of copper pipe, stagnation, corrosion control, and disinfectant residual influenced reduction of le-gionella pneumophila during simulations of the flint water crisis. Pathogens 9, 1–15. https://doi.org/10.3390/pathogens9090730

McGuinness, S.L., O’Toole, J., Barker, S.F., Forbes, A.B., Boving, T.B., Giriyan, A., Patil, K., D’Souza, F., Vhaval, R., Cheng, A.C., Leder, K., 2020. Household Water Storage Management, Hygiene Practices, and Associated Drinking Water Quality in Rural India. Environ. Sci. Technol. 54, 4963–4973. https://doi.org/10.1021/acs.est.9b04818

Muftah, A.M., 2014. 3D fluid flow in an elbow meter-CFD model 1–19.

Pal, C., Bengtsson-Palme, J., Kristiansson, E., Larsson, D.G.J., 2015. Co-occurrence of resistance genes to antibiotics, biocides and metals reveals novel insights into their co-selection potential. BMC Genomics 16, 1–14. https://doi.org/10.1186/s12864-015-2153-5

Rhoads, W.J., Bradley, T.N., Mantha, A., Buttling, L., Keane, T., Pruden, A., Edwards, M.A., 2020. Residential water heater cleaning and occurrence of Legionella in Flint, MI. Water Res. 171, 115439. https://doi.org/10.1016/j.watres.2019.115439

Rhoads, W.J., Hammes, F., 2021. Growth of: Legionella during COVID-19 lockdown stagnation. Environ. Sci. Water Res. Technol. 7, 10–15. https://doi.org/10.1039/d0ew00819b

Richard, R., Boyer, T.H., 2021. Pre-and post-flushing of three schools in Arizona due to COVID-19 shutdown.pdf.

Seiler, C., Berendonk, T.U., 2012. Heavy metal driven co-selection of antibiotic resistance in soil and water bodies impacted by agriculture and aquaculture. Front. Microbiol. 3, 1–10. https://doi.org/10.3389/fmicb.2012.00399

Shaheed, A., Orgill, J., Montgomery, M.A., Jeuland, M.A., Brown, J., 2014. Why “improved” water sources are not always safe. Bull. World Health Organ. 92, 283–289. https://doi.org/10.2471/blt.13.119594

Wang, H., Masters, S., Falkinham, J.O., Edwards, M.A., Pruden, A., 2015. Distribution Sys-tem Water Quality Affects Responses of Opportunistic Pathogen Gene Markers in Household Water Heaters. https://doi.org/10.1021/acs.est.5b01538

World Health Organization, 2017. Guidelines for drinking-water quality, 4th edition: 1st ad-dendum. World Heal. Organ. 1–4.

World Resources Institute, 2019. Aqueduct Water Risk Atlas [WWW Document]. URL https://www.wri.org/applications/aqueduct/water-risk-atlas/#/?advanced=false&base-map=hydro&indicator=w_awr_def_tot_cat&lat=30&lng=-80&map-Mode=view&month=1&opacity=0.5&ponderation=DEF&predefined=false&projec-tion=absolute&scenario=optimistic&scope=baseline&threshold&timeScale=an-nual&year=baseline&zoom=3 (accessed 6.27.22).

Ye, C., Xian, X., Bao, R., Zhang, Y., Feng, M., Lin, W., Yu, X., 2022. Recovery of microbio-logical quality of long-term stagnant tap water in university buildings during the COVID-19 pandemic. Sci. Total Environ. 806, 150616. https://doi.org/10.1016/j.sci-totenv.2021.150616

