## Supplemental Information for "Flushing water regularly inadequate to protect water quality in gravity-fed water supply systems with storage tanks during long-term lockdown"

### SUPPLEMENTARY INFORMATION

**Table S1 The identity of the water samples from four conditions provided in lab scale water distribution network**

| Identity | Description |
|---|---|
| 1D | Tap water with one day stagnation |
| 3D | Tap water with three days stagnation |
| IN | Inlet from the tank |

**Table S2 Primer sequences for the qPCR to target select pathogens and genetic markers**

| Targeted genes/ pathogens* | Primer sequence | References | Analytical LOQ |
|---|---|---|---|

| | | | |
|---|---|---|---|
| | | | **(gene copies/µL of DNA)** |
| 16S rRNA | 5'- CGGTGAATACGTTCYCGG- 3'<br>5'- GGWTACCTTGTTACGACTT- 3' | (Suzuki et al., 2000) | 1000 |
| *sul*1 | 5'- CGCACCGGAAACATCGCTGCAC- 3'<br>5'- TGAAGTTCCGCCGCAAGGCTCG- 3' | (Lee et al., 2011) | 10 |
| *sul*2 | 5'- GAATAAATCGCTCATCATTTTCGG- 3'<br>5'- CGAATTCTTGCGGTTTCTTTCAGC- 3' | (Lee et al., 2011) | 100 |
| *bla*OXA-1 | 5'-CAAGCCAAAGGCACGATAGT- 3'<br>5'-ACGATTGCCTCCCTCTTGAA- 3' | (Wajid et al., 2019) | 10 |
| *erm*F | 5'-CGACACAGCTTTGGTTGAAC-3'<br>5'-GGACCTACCTCATAGACAAG-3' | (Chen et al., 2007) | 10 |
| *int*I1 | 5’-GGGTCAAGGATCTGGATTTCG-3’<br>5’-ACATGCGTGTAAATCATCGTCG-3’ | (Lee et al., 2011) | 10 |
| *L. pneumophila* (*mip*) | 5’-AAAGGCATGCAAGACGCTATG-3’<br>5’-GAAACTTGTTAAGAACGTCTTTCATTTG-3’<br>Probe: FAM-TGGCGCTCAATTGGCTTTAACCGA-BHQ | (Wang et al., 2012) | 10 |
| | 5’-AGAGTTTGATCCTGGCTCAG-3’ | | 10 |

| | | | |
|---|---|---|---|
| *M. avium* (16S rRNA gene) | 5’-ACCAGAAGACATGCGTCTTG-3’ | (Wang et al., 2012) | |
| *E. coli* (*ycc*T) | 5’- GCATCGTGACCACCTTGA-3’<br>5’- CAGCGTGGTGGCAAAA-3’ | (Hembach et al., 2017) | 1000 |

* The annealing temperature for all reactions was 60ºC. The overall qPCR assay contained DNA sample, the forward and reverse primers (500nM), PowerUp™ SYBR™ Green Master Mix (Applied Biosystems, USA) and molecular biology grade water. The total volume of all the qPCR reactions was 7µL and the qPCR was done using the Rotor-Gene Q® (Qiagen®, Hilden, Germany). The amplification efficiency ranged from 80% to 110% and the correlation coefficient of the standard curve 0.95 to 0.99. every run included molecular biology grade water as no-template control.

**Table S3 Correlation of heavy metals i.e., iron, lead, copper and zinc and other water physicochemical parameters i.e., conductivity, pH, TOC, chloride, nitrate, and sulphate in tap water collected from laboratory-scale simulation of premise plumbing subjected to one day stagnation without contamination (*: P-value < 0.05, **: P-value < 0.01, ***: P-value < 0.001)**

| | 16S rRNA | *sul*1 | *sul*1/ bacterium | *int*I1 | *int*I1/ bacterium | *mip* | *mip*/ bacterium | Iron | Lead | Copper | Zinc | Total organic carbon | Conductivity | pH | Chloride | Nitrate | Sulphate |
|---|---|---|---|---|---|---|---|---|---|---|---|---|---|---|---|---|---|
| *sul*1 | 0.62**** | | | | | | | | | | | | | | | | |
| *sul*1/ bacterium | -0.37*** | 0.37*** | | | | | | | | | | | | | | | |
| *int*I1 | 0.70**** | 0.53**** | -0.1 | | | | | | | | | | | | | | |
| *int*I1/ bacterium | 0.45**** | 0.40**** | -0.01 | 0.89**** | | | | | | | | | | | | | |
| *mip* | 0.22* | 0.14 | -0.1 | 0.18 | 0.11 | | | | | | | | | | | | |
| *mip*/bacterium | -0.52**** | -0.28** | 0.22* | -0.30** | -0.18 | 0.58**** | | | | | | | | | | | |
| Iron | 0.25* | 0.14 | -0.05 | 0.16 | 0.04 | 0.30** | 0.05 | | | | | | | | | | |
| Lead | 0.13 | -0.05 | -0.2 | -0.05 | -0.15 | 0.35*** | 0.21* | 0.81**** | | | | | | | | | |
| Copper | 0.05 | -0.1 | -0.19 | -0.1 | -0.14 | 0.27** | 0.11 | 0.55**** | 0.69**** | | | | | | | | |
| Zinc | 0.16 | -0.03 | -0.17 | 0.06 | -0.01 | 0.32** | 0.15 | 0.70**** | 0.85**** | 0.68**** | | | | | | | |
| Total organic carbon | 0.17 | 0.07 | -0.22* | -0.01 | -0.05 | 0.05 | -0.01 | 0.28** | 0.40**** | 0.33** | 0.26* | | | | | | |
| Conductivity | 0.13 | 0.02 | -0.03 | 0.14 | 0.04 | 0.01 | -0.06 | 0.19 | 0.15 | 0.06 | 0.13 | 0.05 | | | | | |
| pH | 0.05 | -0.08 | -0.11 | -0.12 | -0.21* | 0.16 | 0.15 | 0.42**** | 0.47**** | 0.35*** | 0.38*** | 0.44**** | 0.15 | | | | |
| Chloride | 0.15 | 0.16 | 0.03 | 0.31** | 0.30** | 0.01 | -0.09 | -0.03 | -0.25* | -0.39*** | -0.17 | -0.01 | 0.11 | -0.09 | | | |
| Nitrate | -0.06 | 0.09 | 0.17 | 0.15 | 0.28** | -0.1 | -0.01 | 0.07 | -0.13 | -0.2 | -0.01 | -0.15 | 0 | -0.05 | 0.27** | | |
| Sulphate | 0.17 | 0.15 | -0.05 | 0.1 | 0.02 | -0.07 | -0.15 | -0.23* | -0.35*** | -0.33** | -0.49**** | -0.06 | -0.30** | -0.12 | 0.33** | 0 | |
| Chloride to Sulphate Mass Ratio | -0.12 | -0.03 | 0.12 | 0.05 | 0.12 | 0.07 | 0.12 | 0.15 | 0.14 | 0.03 | 0.29** | 0.01 | 0.34*** | 0.05 | 0.36*** | 0.07 | -0.70**** |

**Table S4 Correlation of heavy metals i.e., iron, lead, copper and zinc and other water physicochemical parameters i.e., conductivity, pH, TOC, chloride, nitrate, and sulphate in tap water collected from laboratory-scale simulation of premise plumbing subjected to three days stagnation without contamination (*: P-value < 0.05, **: P-value < 0.01, ***: P-value < 0.001)**

| | 16S rRNA | *sul*1 | *sul*1/ bacterium | *int*I1 | *int*I1/ bacterium | *mip* | *mip*/ bacterium | Iron | Lead | Copper | Zinc | Total organic carbon | Conductivity | pH | Chloride | Nitrate | Sulphate |
|---|---|---|---|---|---|---|---|---|---|---|---|---|---|---|---|---|---|
| *sul*1 | 0.53**** | | | | | | | | | | | | | | | | |
| *sul*1/ bacterium | -0.68**** | 0.14 | | | | | | | | | | | | | | | |
| *int*I1 | 0.69**** | 0.56**** | -0.35*** | | | | | | | | | | | | | | |
| *int*I1/ bacterium | 0.04 | 0.29** | 0.13 | 0.72**** | | | | | | | | | | | | | |
| *mip* | 0.14 | -0.04 | -0.2 | 0.01 | -0.13 | | | | | | | | | | | | |
| *mip*/ bacterium | -0.29** | -0.23* | 0.08 | -0.25* | -0.11 | 0.83**** | | | | | | | | | | | |
| Iron | 0.08 | 0.21* | 0.05 | 0.01 | -0.05 | 0.29** | 0.24* | | | | | | | | | | |
| Lead | -0.07 | -0.04 | 0.06 | -0.17 | -0.14 | 0.24* | 0.22* | 0.81**** | | | | | | | | | |
| Copper | 0.09 | -0.01 | -0.06 | 0.05 | 0.02 | 0.15 | 0.12 | 0.61**** | 0.70**** | | | | | | | | |
| Zinc | -0.13 | -0.16 | 0.03 | -0.21* | -0.15 | 0.28** | 0.27** | 0.66**** | 0.82**** | 0.60**** | | | | | | | |
| Total organic carbon | -0.07 | -0.16 | -0.02 | -0.11 | -0.1 | -0.04 | -0.03 | 0.23* | 0.44**** | 0.38*** | 0.43**** | | | | | | |
| Conductivity | 0.04 | 0.15 | 0.04 | 0 | -0.04 | 0.1 | 0.06 | -0.21* | -0.27** | -0.46**** | -0.21* | -0.19 | | | | | |
| pH | 0.08 | 0.01 | -0.05 | 0 | -0.09 | 0.05 | -0.03 | 0.13 | 0.13 | 0.11 | 0.24* | 0.15 | 0.17 | | | | |
| Chloride | 0.30** | 0.25* | -0.16 | 0.21* | -0.07 | -0.04 | -0.1 | -0.13 | -0.31** | -0.34*** | -0.32** | -0.01 | 0.24* | 0.03 | | | |
| Nitrate | -0.11 | 0.03 | 0.08 | -0.12 | -0.09 | -0.07 | 0.05 | -0.1 | -0.21* | -0.31** | -0.17 | -0.07 | 0.22* | 0.04 | 0.27** | | |
| Sulphate | 0.30** | 0.32** | -0.08 | 0.29** | 0.08 | -0.30** | -0.34*** | -0.24* | -0.40**** | -0.18 | -0.45**** | -0.08 | -0.05 | 0.12 | 0.59**** | 0.25* | |
| Chloride to Sulphate Mass Ratio | -0.11 | -0.16 | -0.04 | -0.16 | -0.15 | 0.34*** | 0.33** | 0.11 | 0.19 | -0.08 | 0.22* | 0.05 | 0.22* | -0.2 | 0.08 | -0.18 | -0.68**** |

Table S5: Total organic carbon supplied by PVC storage tank

| | Influent TOC |
|---|---|
| IN_1 | 5.3484 |
| IN_2 | 6.24 |
| IN_3 | 4.7268 |
| IN_4 | 4.308 |
| IN_5 | 9.6 |
| IN_6 | 4.4958 |
| IN_7 | 4.056 |
| IN_8 | 4.7862 |
| IN_9 | 5.772 |
| IN_10 | 6.54 |
| IN_11 | 5.1132 |
| IN_12 | 4.2408 |
| IN_13 | 4.6638 |
| IN_14 | 4.0116 |
| IN_15 | 5.8704 |
| IN_16 | 4.4298 |
| IN_17 | 3.9102 |
| IN_18 | 6.24 |
| IN_19 | 5.0214 |
| IN_20 | 4.1082 |
| IN_21 | 5.5554 |
| IN_22 | 5.7984 |
| IN_23 | 7.5 |
| IN_24 | 6.18 |
| IN_25 | 4.8996 |
| IN_26 | 6.3 |
| IN_27 | 4.8162 |
| IN_28 | 6.9 |
| IN_29 | 5.715 |
| IN_30 | 4.8066 |
| IN_31 | 3.48 |

Table S6: Total organic carbon supplied by OHTs

| | Total Organic Carbon | |
|---|---|---|
| | Monsoon | Winter |
| OHT1 | 4.265 | 10.15 |
| OHT2 | 7.95 | 10.5 |
| OHT3 | 7.15 | 10.05 |
| OHT4 | 5.9 | 9.35 |
| OHT5 | 7.35 | 12.35 |
| OHT6 | 5.95 | 11 |

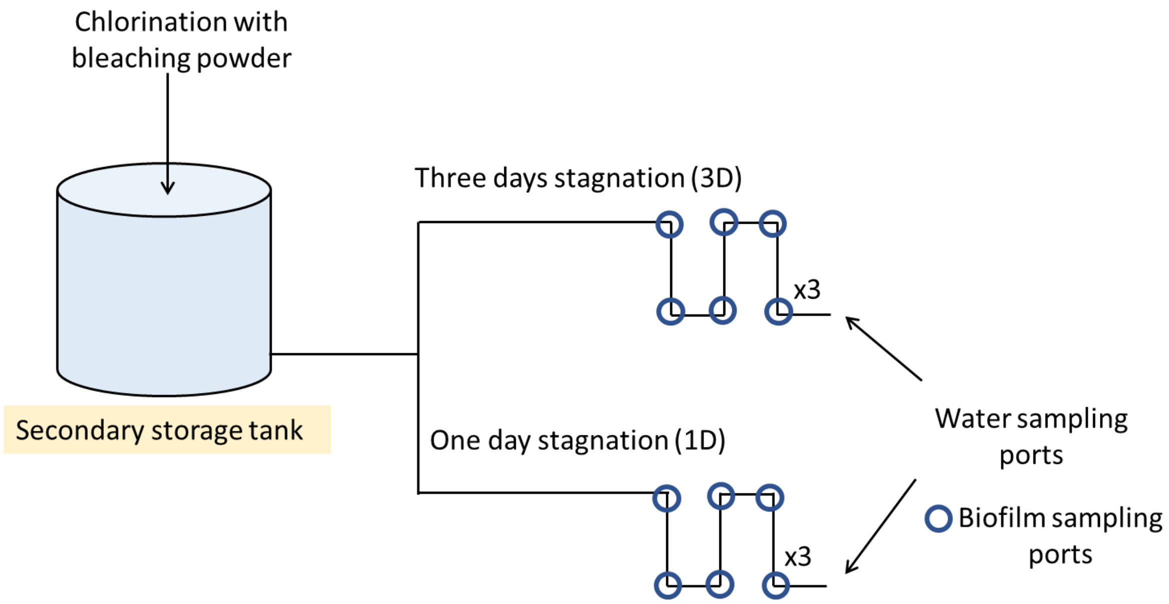


**Figure.S1** Schematic for Lab scale simulated water distribution network with GI pipes

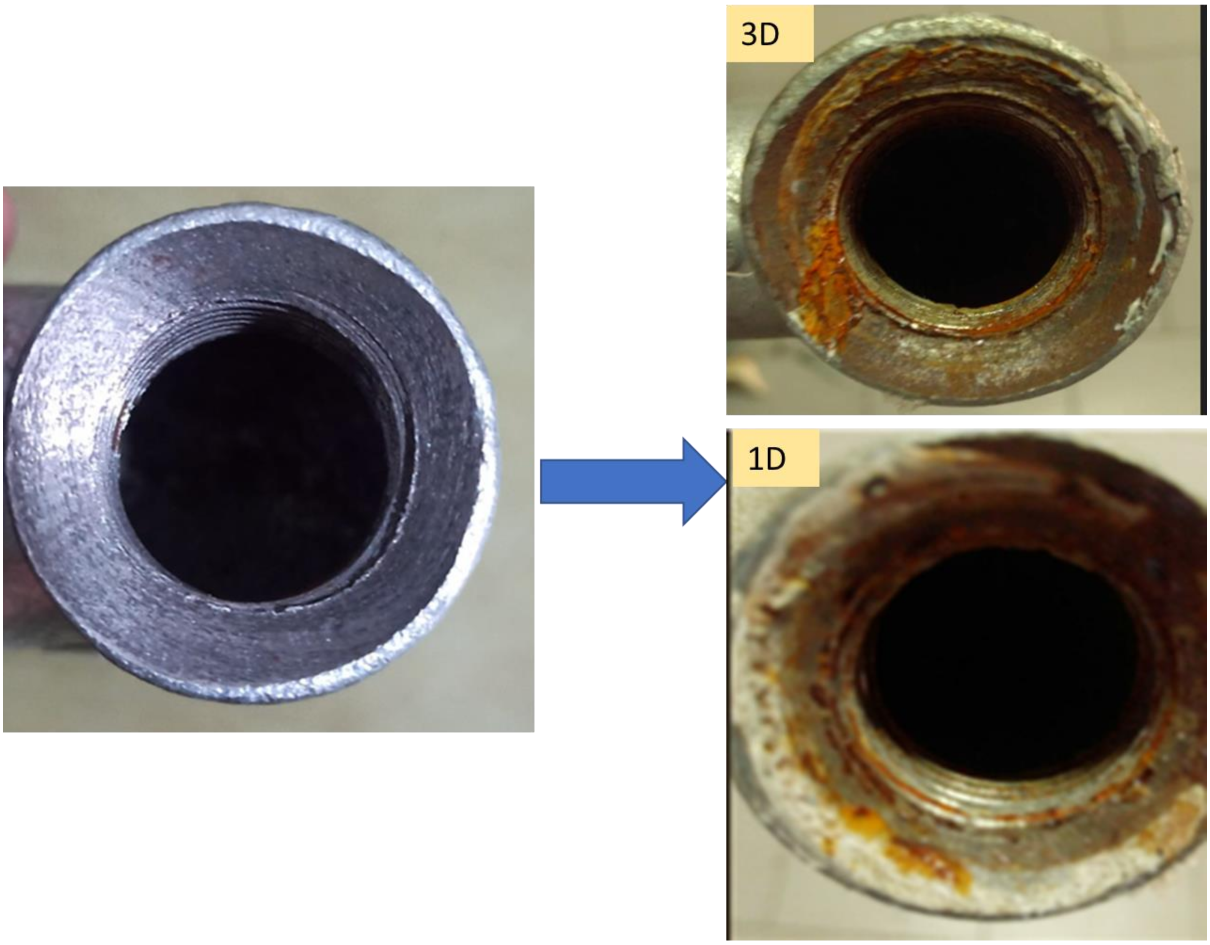


**Figure S2.** Morphological changes in pipes after 52 weeks of installation

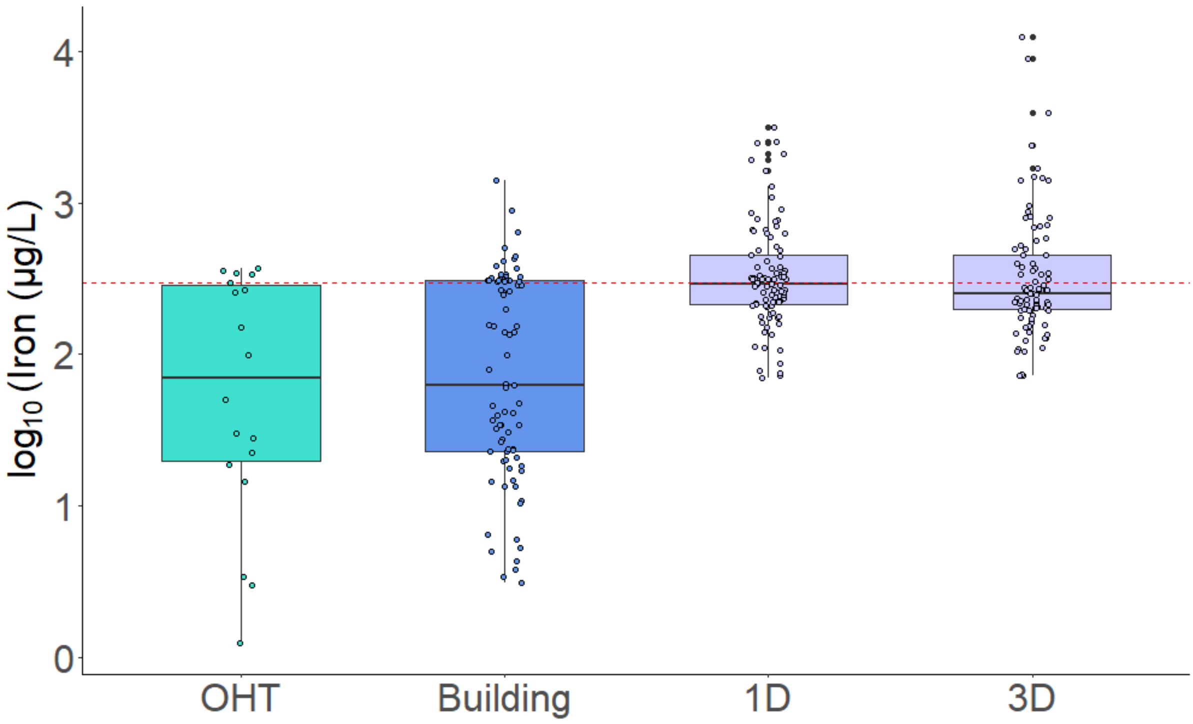


**Figure S3.** The concentration of iron (μg/L) in water samples for OHT and building samples and three different primary branches of laboratory-scale premise water supply network (1D, and 3D). The Red dashed line represents the WHO (World Health Organisation, 2017) and BIS (Bureau of Indian Standards, 2012) guidelines for heavy metal levels in drinking water, respectively.

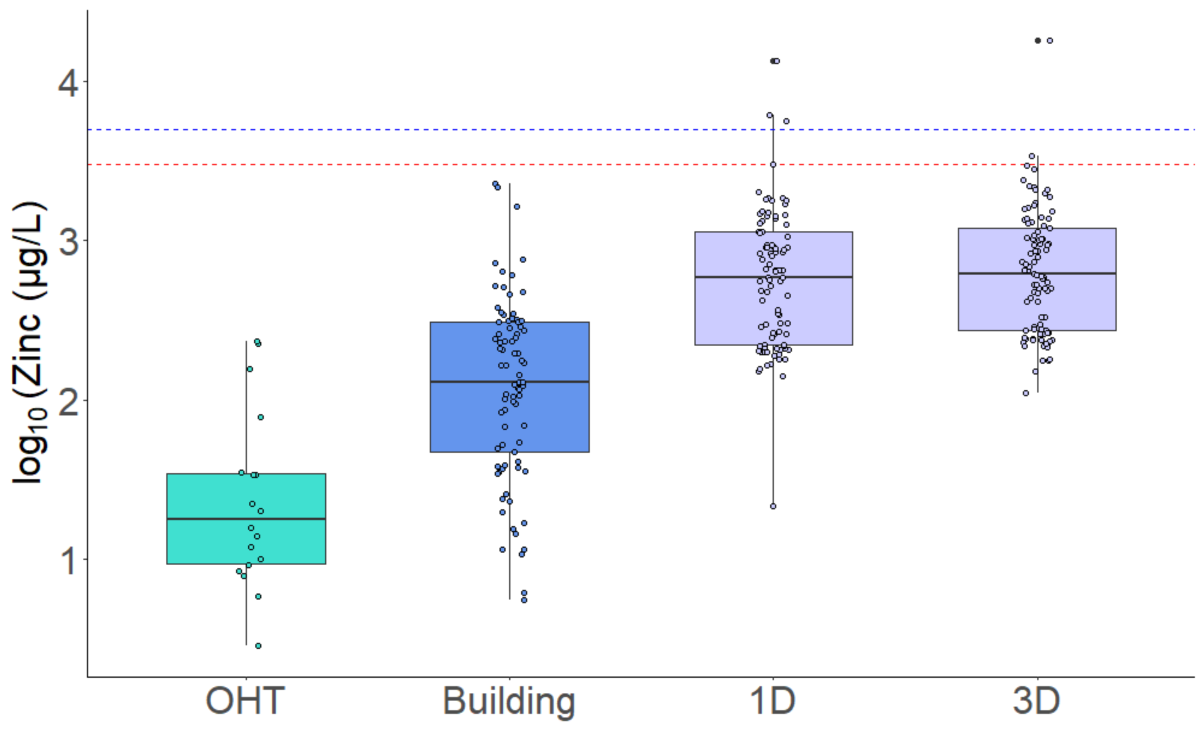


**Figure S4.** The concentration of zinc (µg/L) in water samples for OHT and building samples and three different primary branches of laboratory-scale premise water supply network (1D, and 3D). Red and blue dashed lines represent the WHO and BIS guidelines for heavy metal levels in drinking water, respectively.

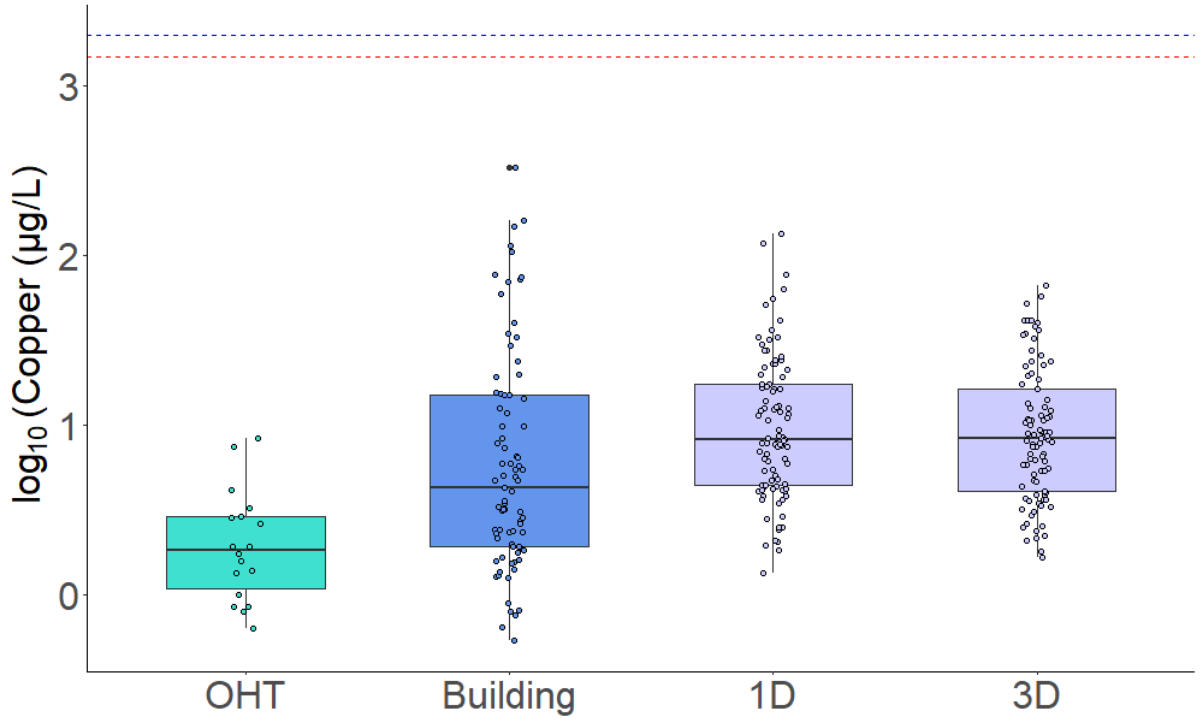


**Figure S5.** The concentration of copper (µg/L) in water samples for OHT and building samples and three different primary branches of laboratory-scale premise water supply network (1D, and 3D). Red and blue dashed lines represent the WHO (World Health Organisation, 2017) and BIS (Bureau of Indian Standards, 2012) guidelines for heavy metal levels in drinking water, respectively.

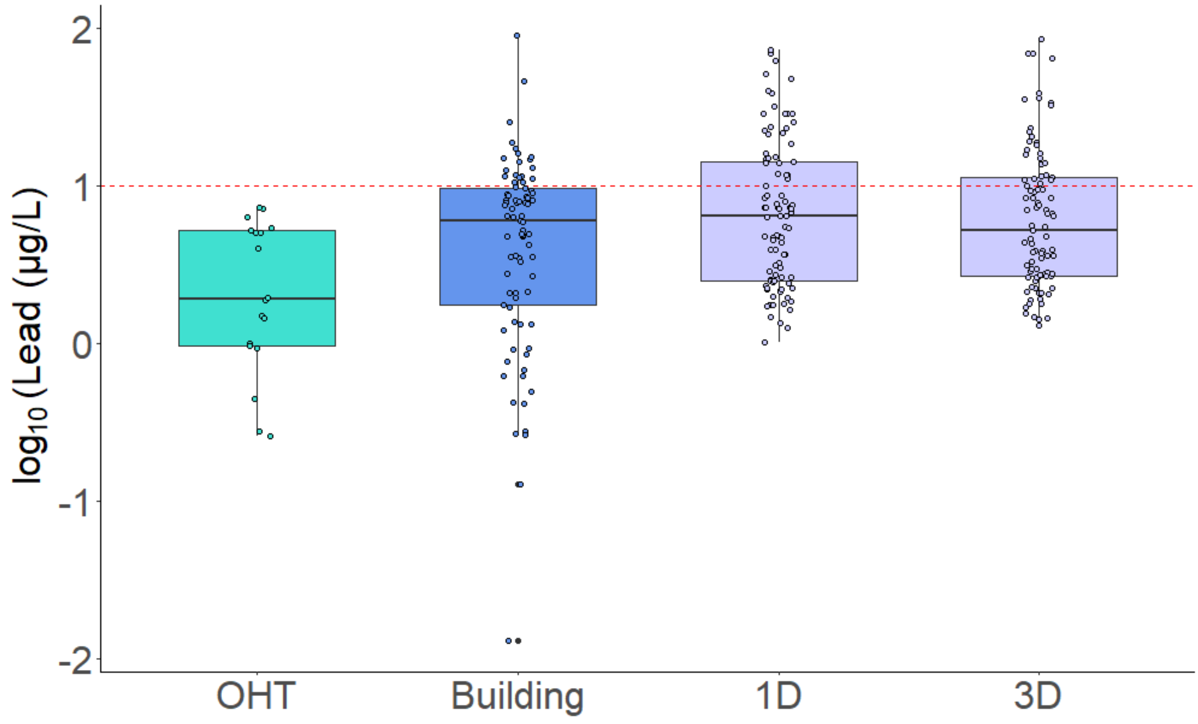


**Figure S6** The concentration of lead (μg/L) in water samples for OHT and building samples and three different primary branches of laboratory-scale premise water supply network (*1*D, *and* 3D). The Red dashed line represents the WHO (World Health Organisation, 2017) guidelines for heavy metal levels in drinking water, respectively.

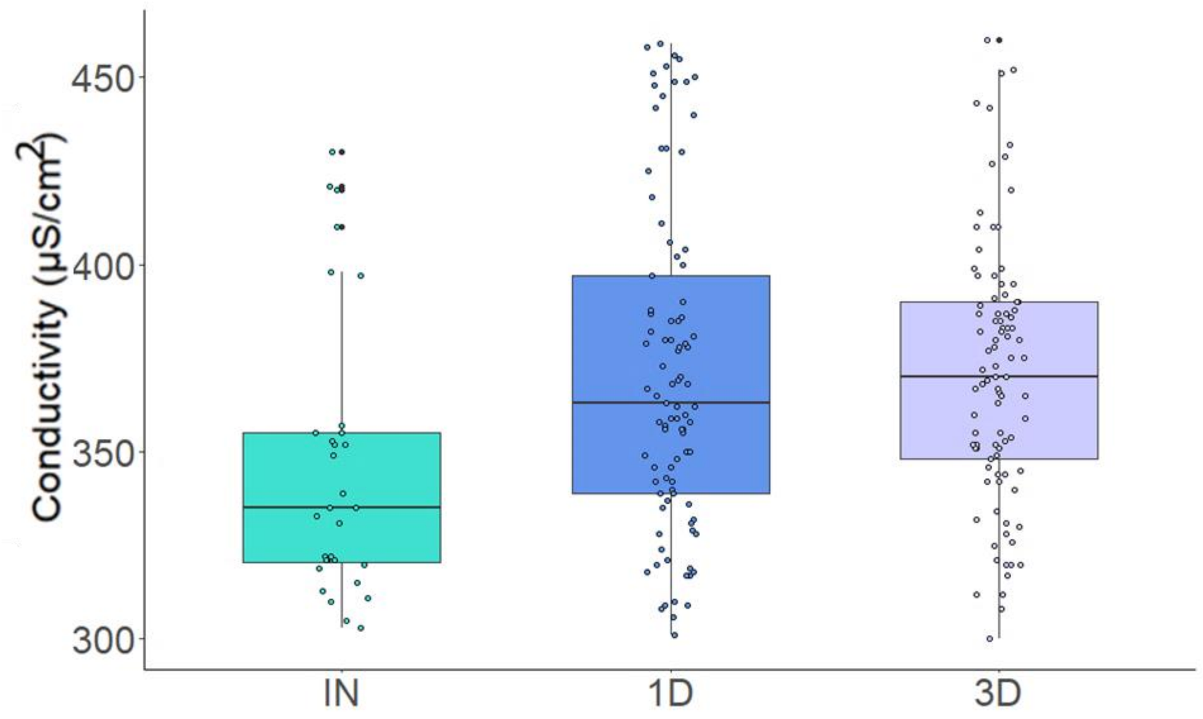


**Figure S7.** The conductivity (µS/cm$^2$) values in water samples resolved across the two different conditions (1D, and 3D). IN represents the influent concentration.

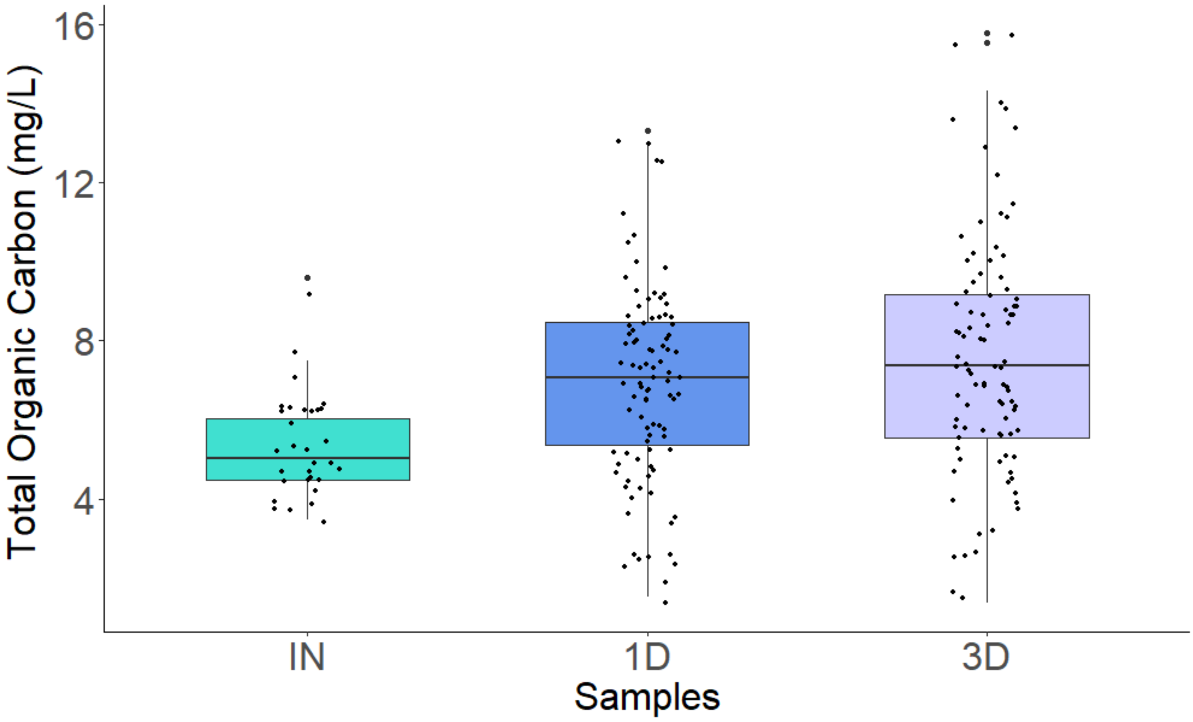


**Figure S8.** The values of total organic carbon (mg/L) in water samples resolved across the two different conditions (1D, and 3D). IN represents the influent concentration.

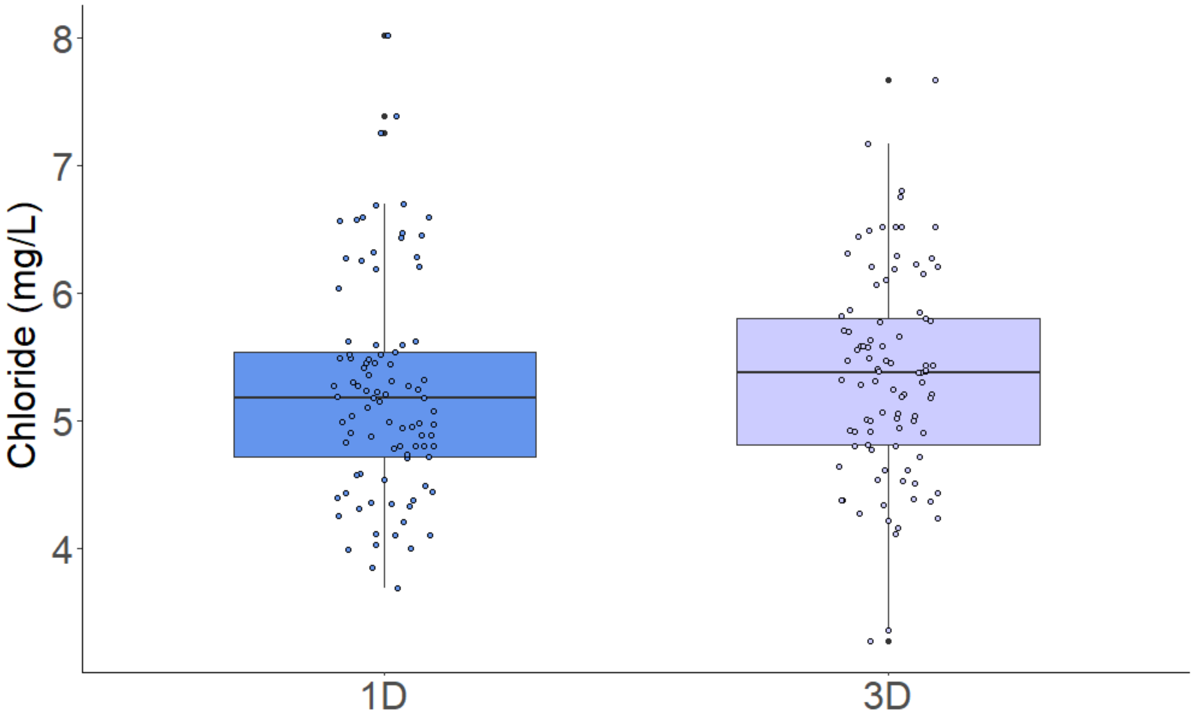


**Figure S9** The chloride (mg/L) values in water samples resolved across the two different conditions (1D, and 3D).

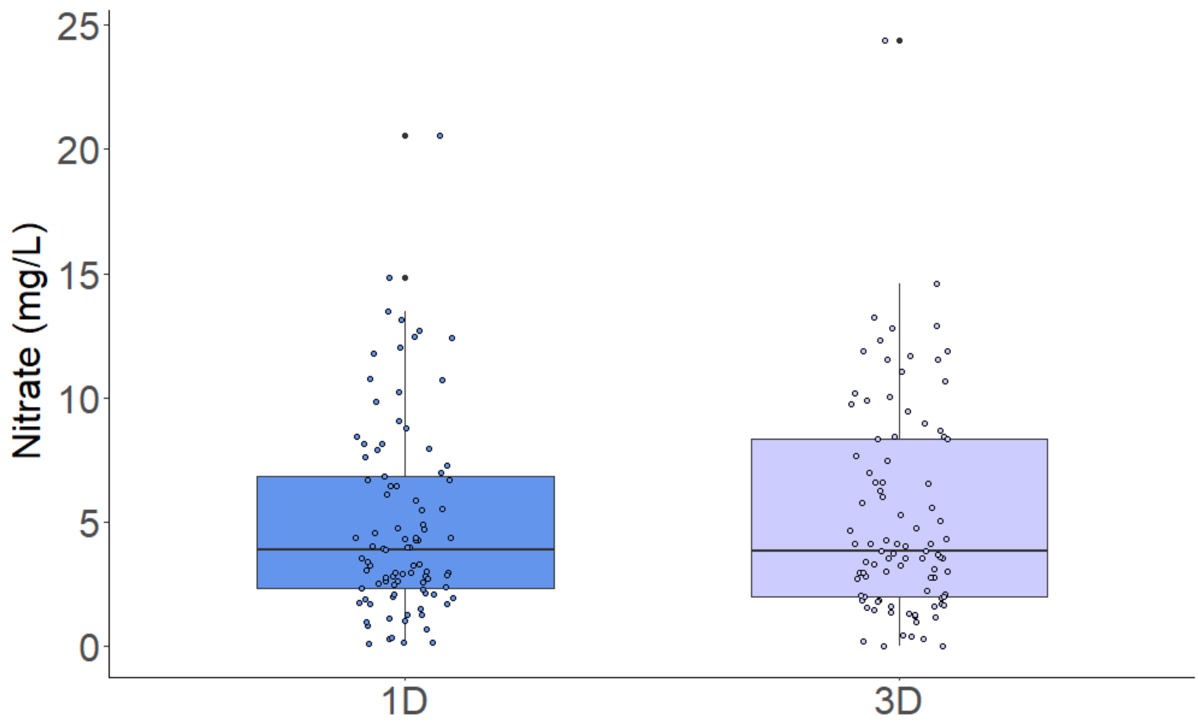


**Figure S10.** The nitrate (mg/L) values in water samples resolved across the two different conditions (1D, and 3D).

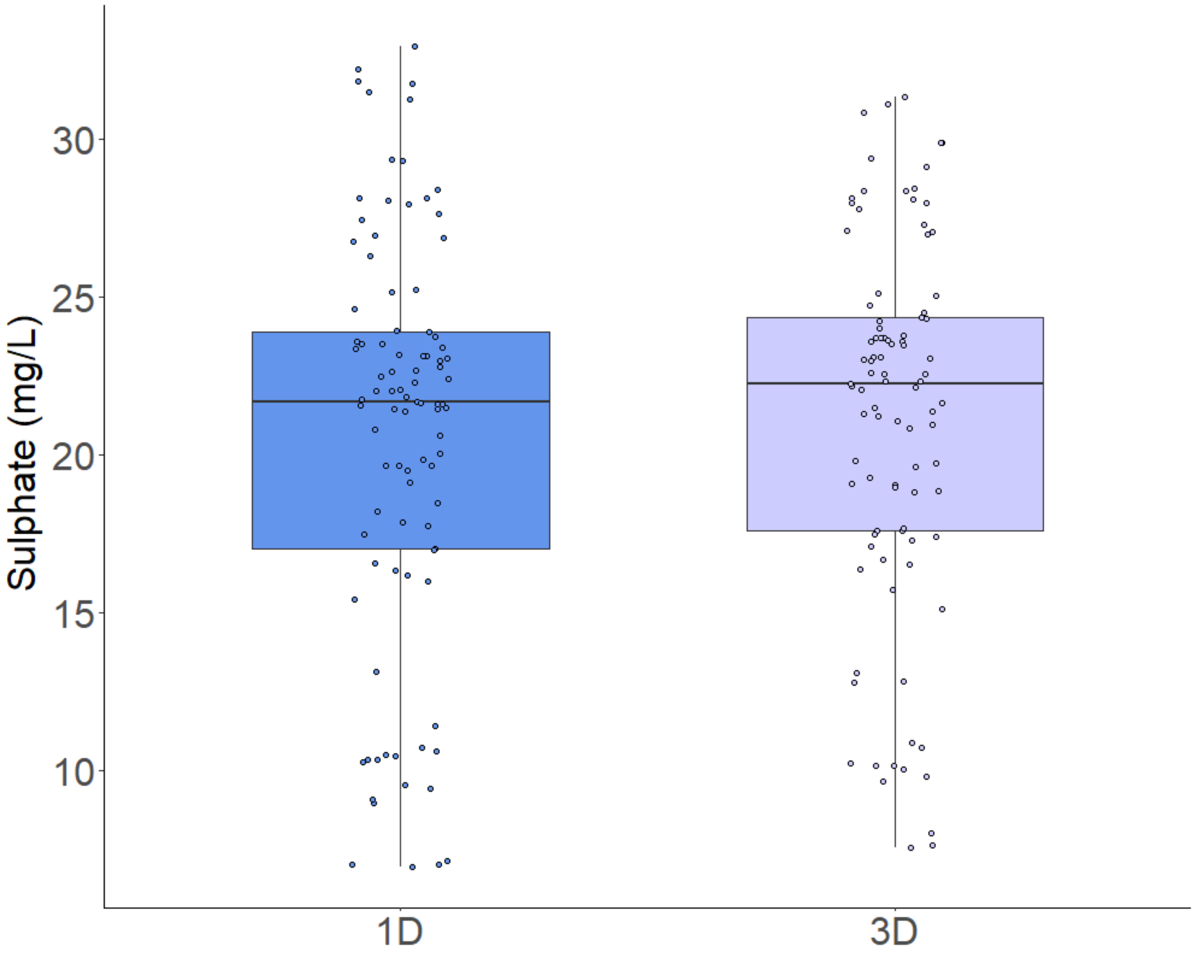


**Figure S11.** The sulphate (mg/L) values in water samples resolved across the two different conditions (1D, and 3D).

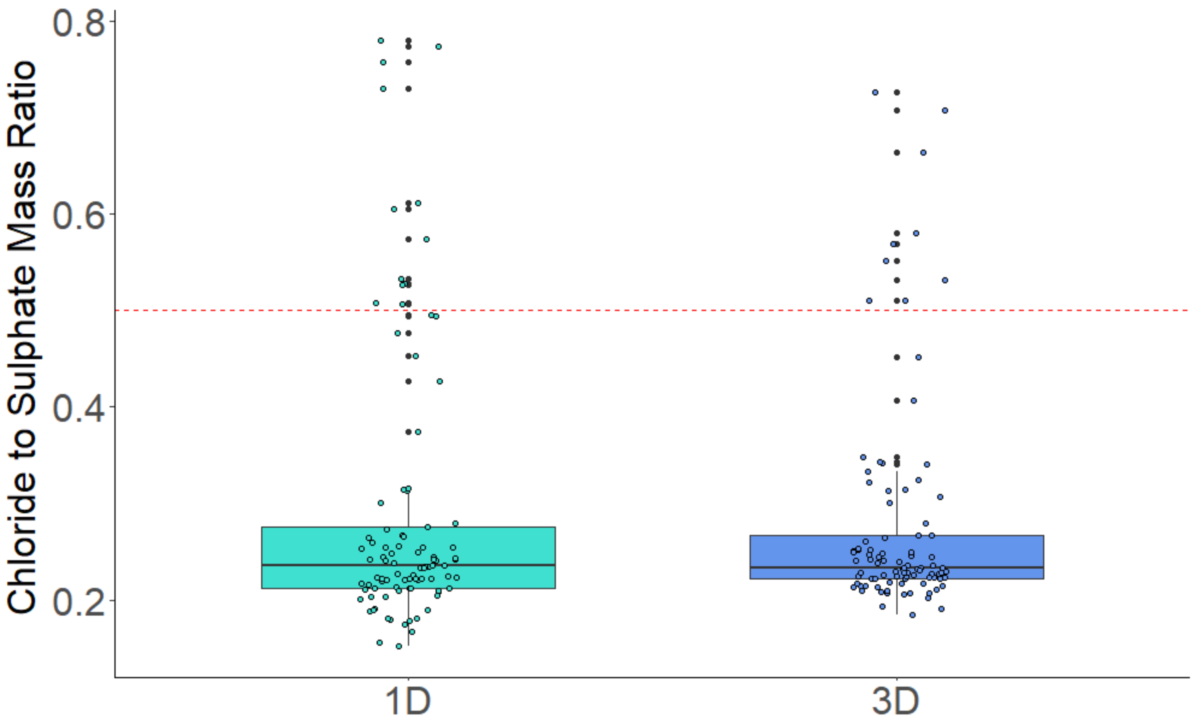


**Figure S12.** The level of Chloride to sulphate mass ratio in water samples for two different conditions (1D, and 3D).

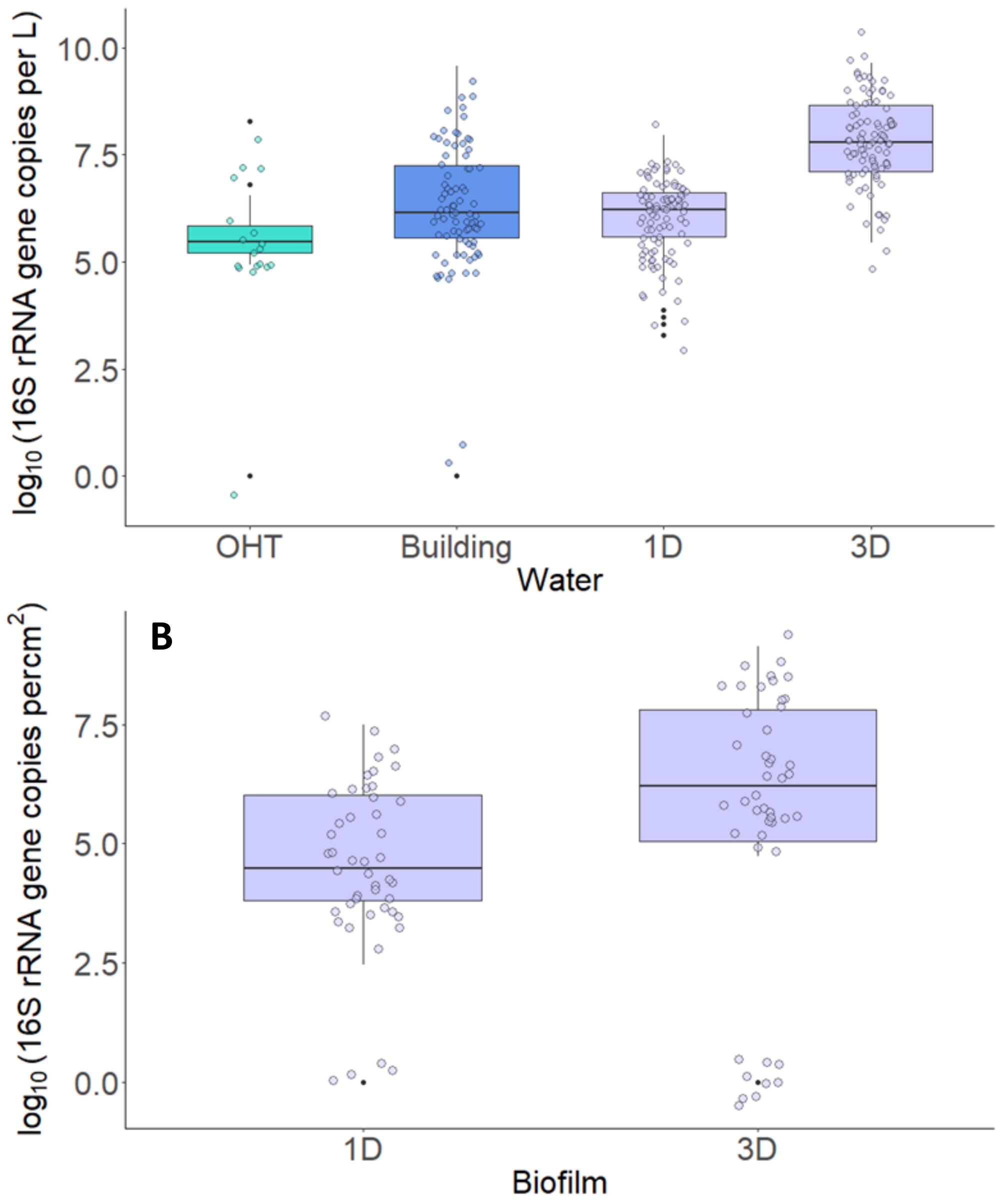


**Figure S13. Total bacterial count.** The y-axis represents levels of total bacterial count as determined by the levels of 16S rRNA gene copies in water samples. The samples in **A.** were collected from the OHTs and the buildings under regular use an year prior to pandemic, and **B.** biofilm samples laboratory-scale simulation of premise plumbing subject to water being drawn once per day (1D) and once in three days (3D) respectively.

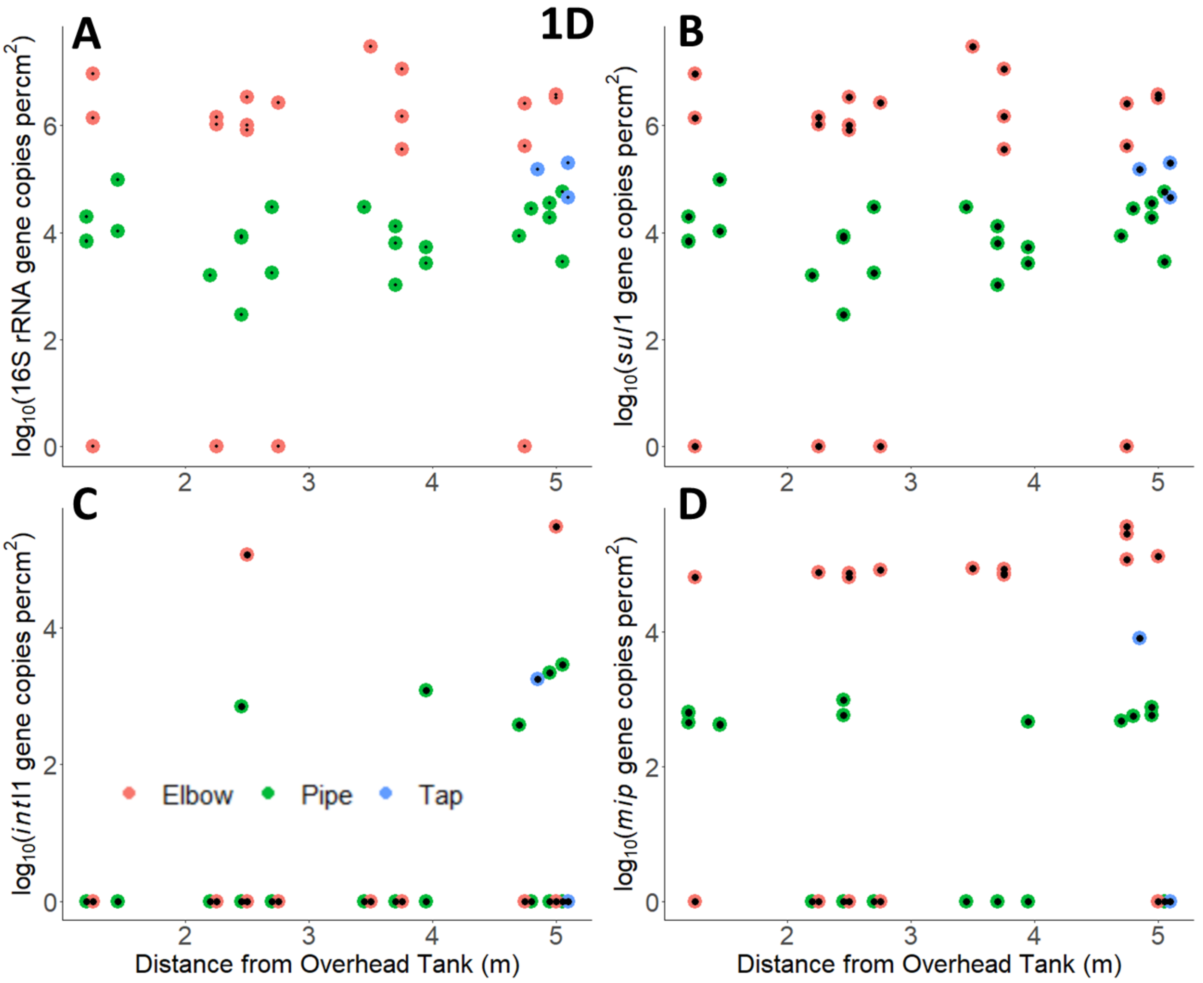


**Figure S14.** The levels of 16S rRNA, *sul*1, *int*I1, and *mip* genes in pipe, elbow and tap biofilm samples in pipes with respect to the distance from overhead storage tank subjected 1D condition refer to water drawn once per day.

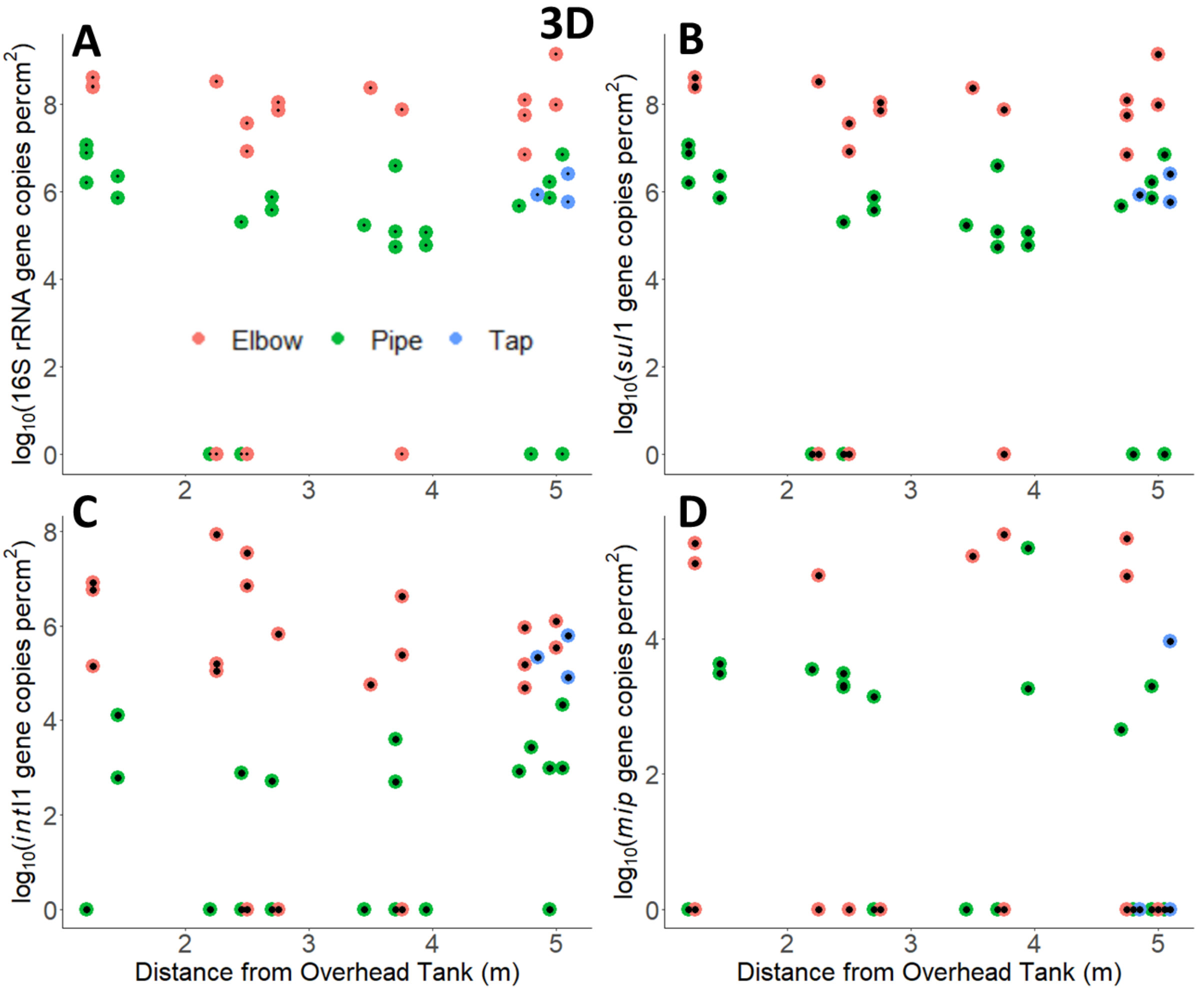


**Figure S15.** The levels of 16S rRNA, *sul*1, *int*I1, and *mip* genes in pipe, elbow and tap biofilm samples in pipes with respect to the distance from overhead storage tank subjected 3D condition refer to water drawn once per day.

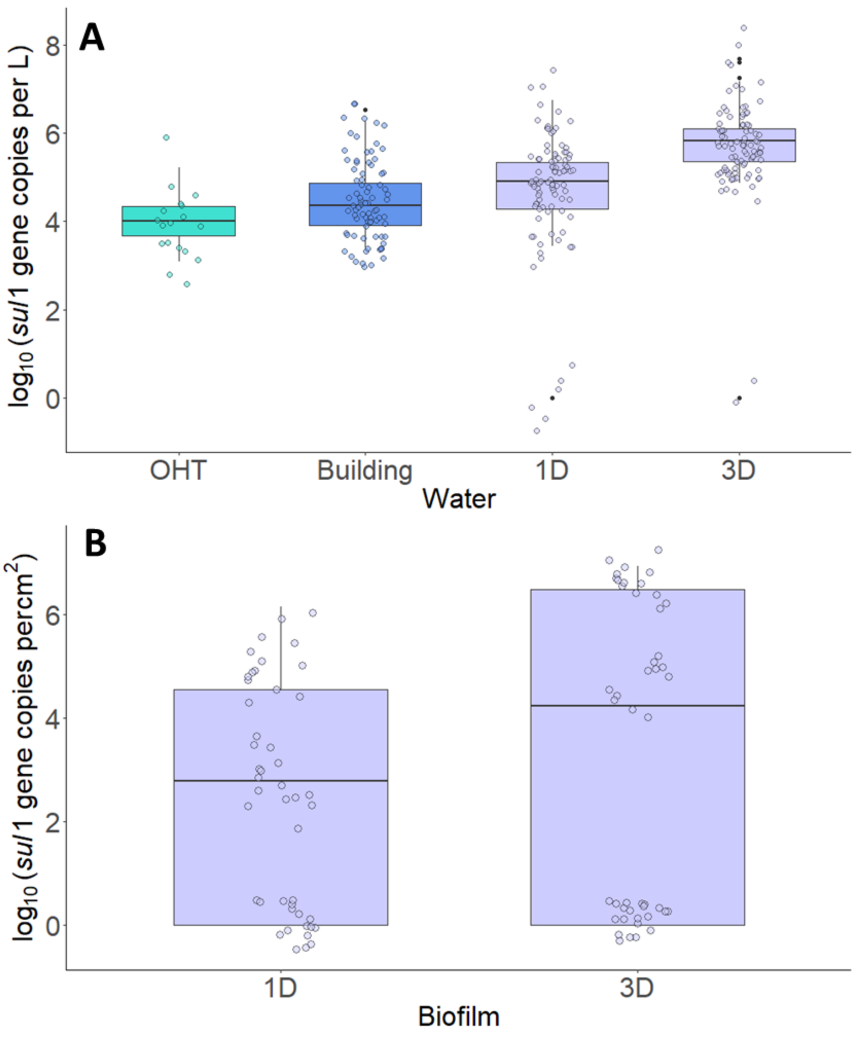


**Figure S16. *sul*1 count.** The y-axis represents levels of *sul*1 gene copies in water samples. The samples in **A.** were collected from the OHTs and the buildings under regular use an year prior to pandemic, and **B.** biofilm samples laboratory-scale simulation of premise plumbing subject to water being drawn once per day (1D) and once in three days (3D) respectively.

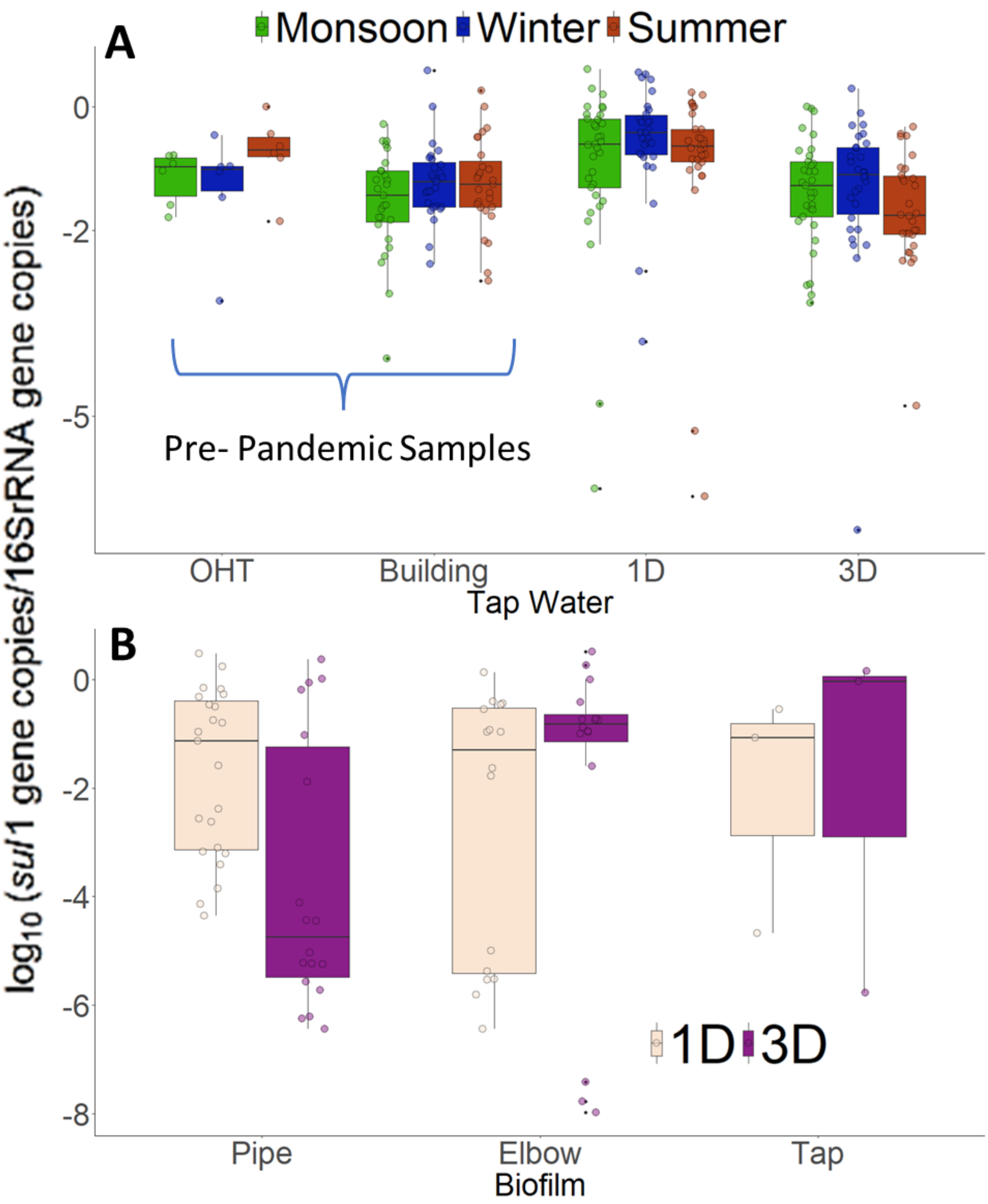


**Figure S17. Levels of *sul*1 gene per bacterium.** The y-axis represents the level of *sul*1 gene copies normalized to total bacterial count, assuming 4.2 16S rRNA gene copies per bacteria. **A.** The tap water from the OHTs and the buildings an year before the pandemic, and from the laboratory-scale water supply network in three different seasons (monsoon, winter, and summer) and **B.** biofilm samples subject to water being drawn once per day (1D) and once in three days (3D) respectively.

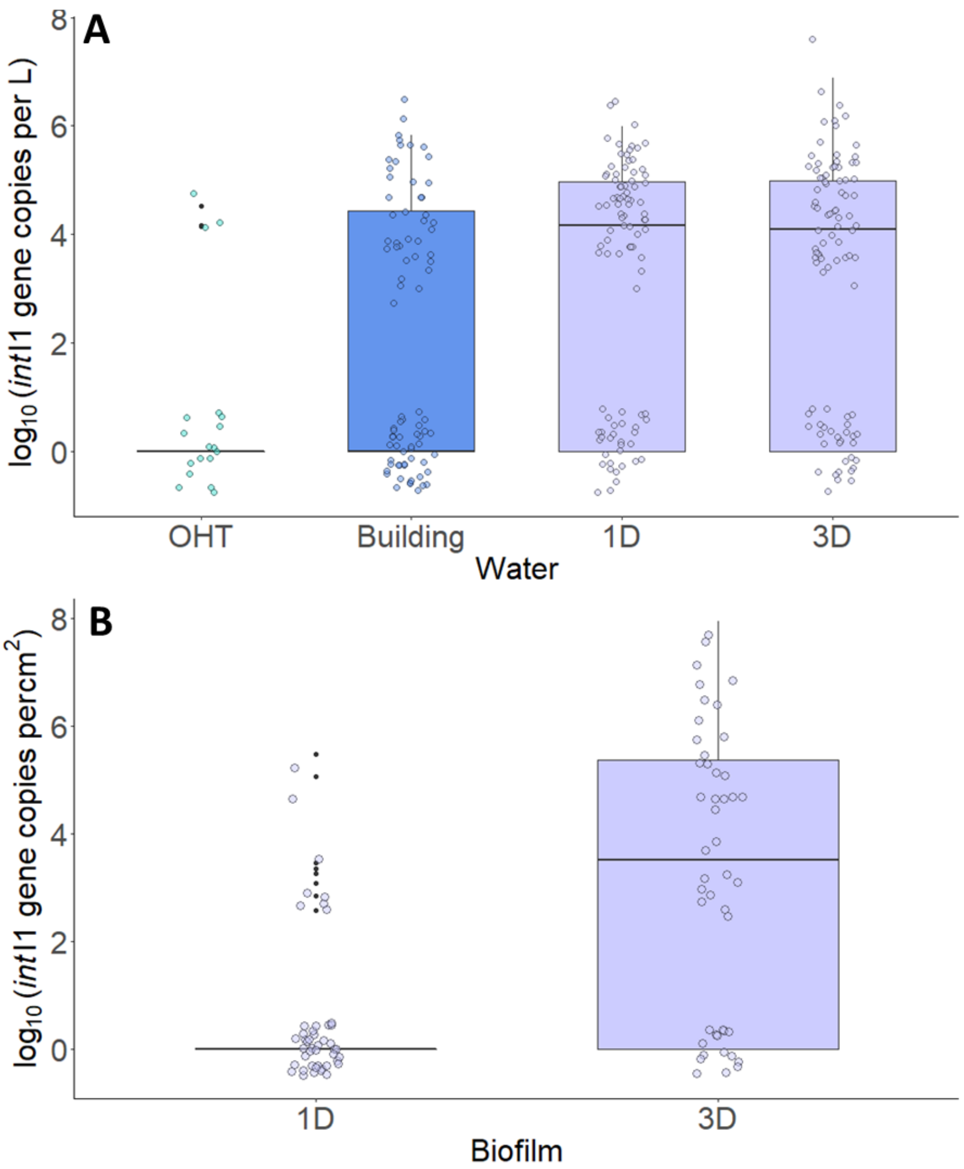


**Figure S18. *int*I1 count.** The y-axis represents levels of *int*I1 gene copies in water samples. The samples in **A.** were collected from the OHTs and the buildings under regular use an year prior to pandemic, and **B.** biofilm samples laboratory-scale simulation of premise plumbing subject to water being drawn once per day (1D) and once in three days (3D) respectively.

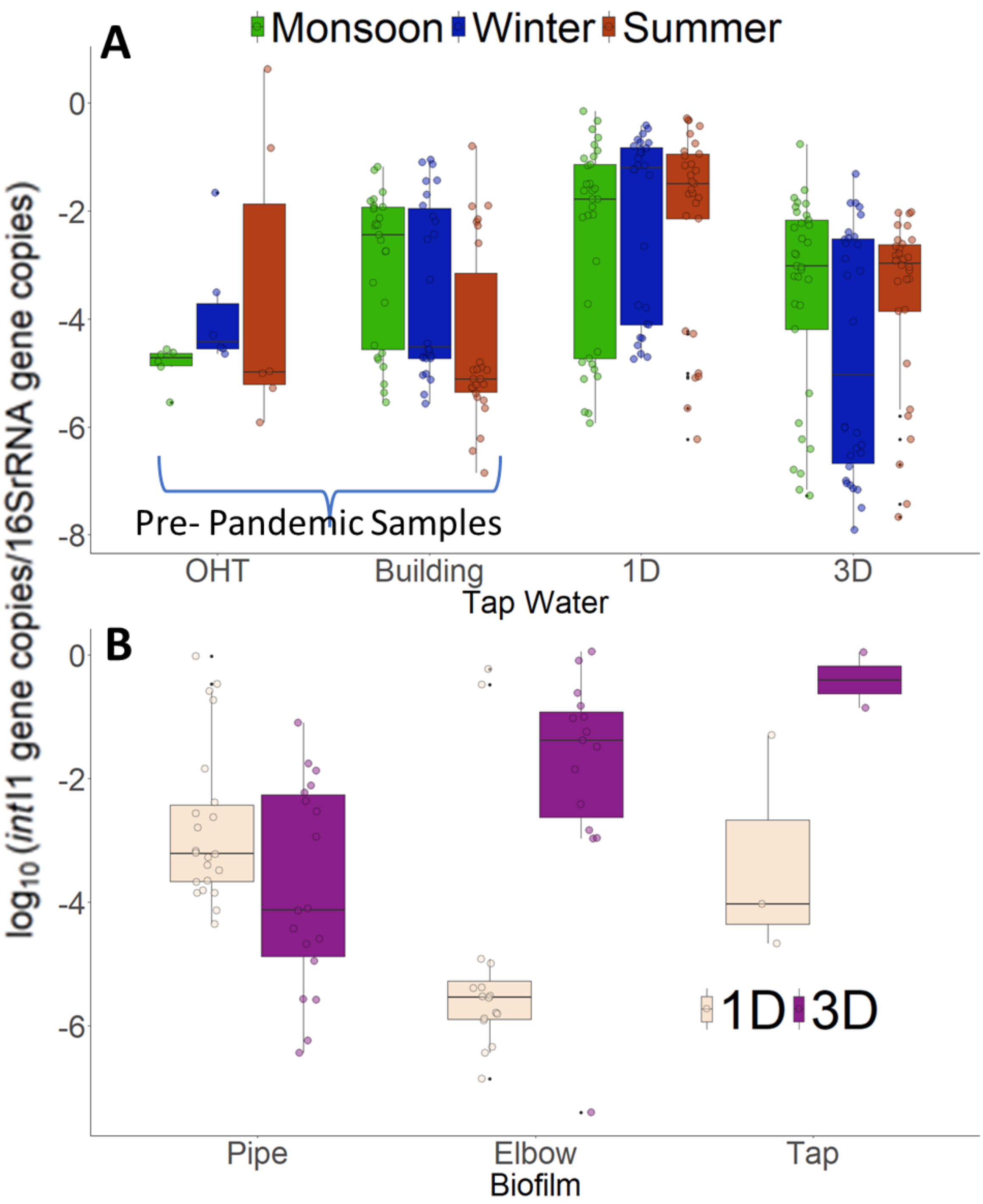


**Figure S19. Levels of *int*I1 gene per bacterium.** The y-axis represents the level of *int*I1 gene copies normalized to total bacterial count, assuming 4.2 16S rRNA gene copies per bacteria. **A.** The tap water from the OHTs and the buildings an year before the pandemic, and from the laboratory-scale water supply network in three different seasons (monsoon, winter, and summer) and **B.** biofilm samples subject to water being drawn once per day (1D) and once in three days (3D) respectively.

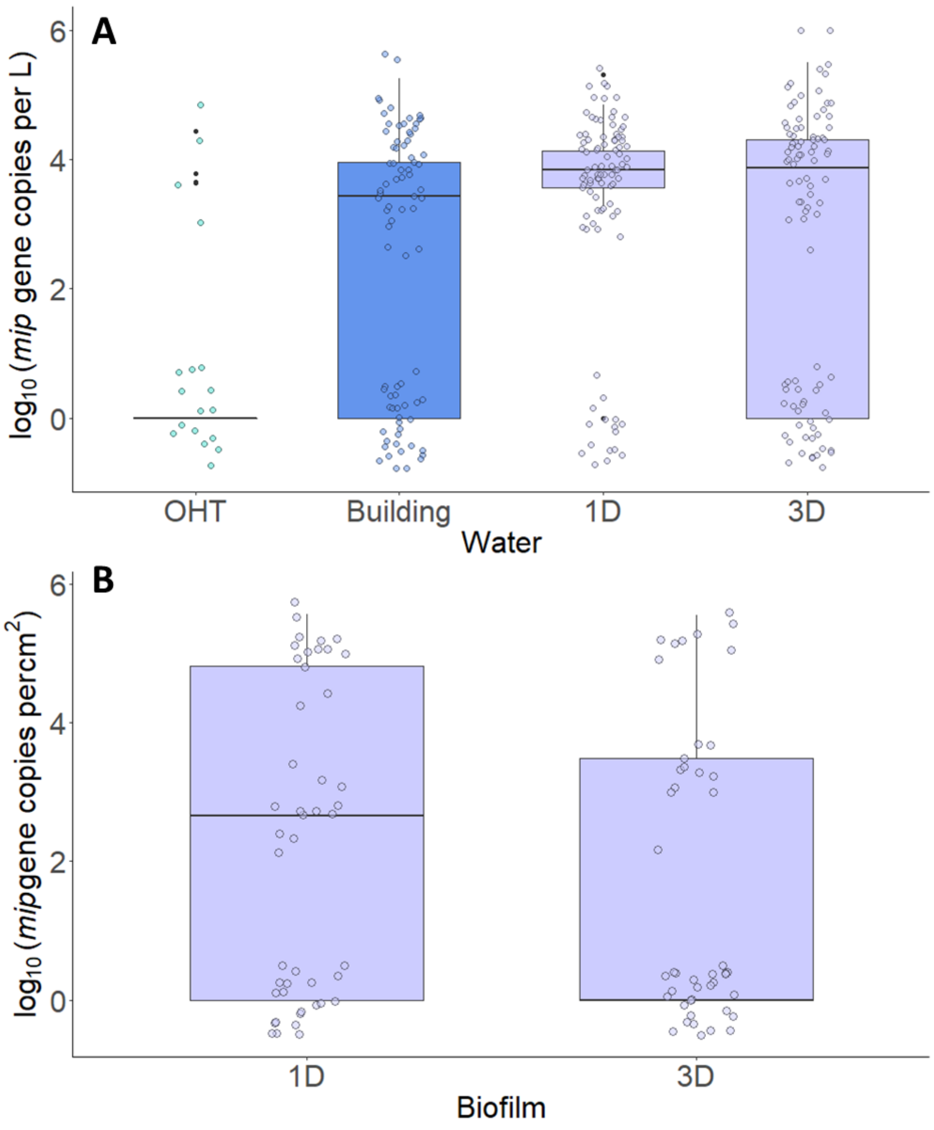


**Figure S20.** ***L. pneumophila (mip)*** **count.** The y-axis represents levels of *mip* gene copies in water samples. The samples in **A.** were collected from the OHTs and the buildings under regular use an year prior to pandemic, and **B.** biofilm samples laboratory-scale simulation of premise plumbing subject to water being drawn once per day (1D) and once in three days (3D) respectively.

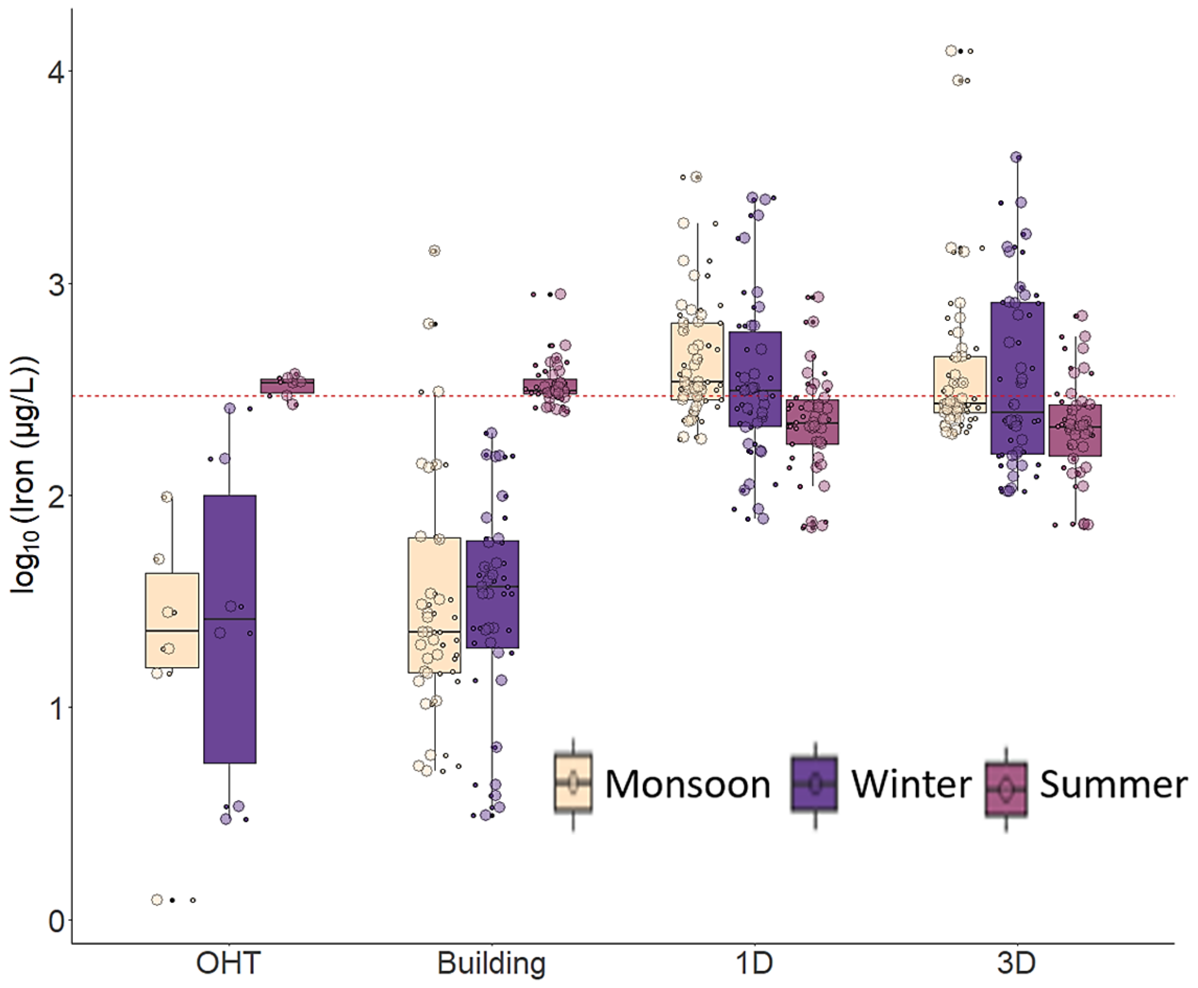


**Figure S8. Levels of Iron (µg/L) in tap water.** The y-axis represents the level of iron (µg/L) in the tap water collected from the OHTs and the buildings an year before the pandemic, and from the laboratory-scale water supply network in three different seasons (monsoon, winter, and summer) subject to water being drawn once per day (1D) and once in three days (3D) respectively. The Red dashed line represents the WHO (World Health Organisation, 2017) and BIS (Bureau of Indian Standards, 2012) guidelines for heavy metal levels in drinking water, respectively.

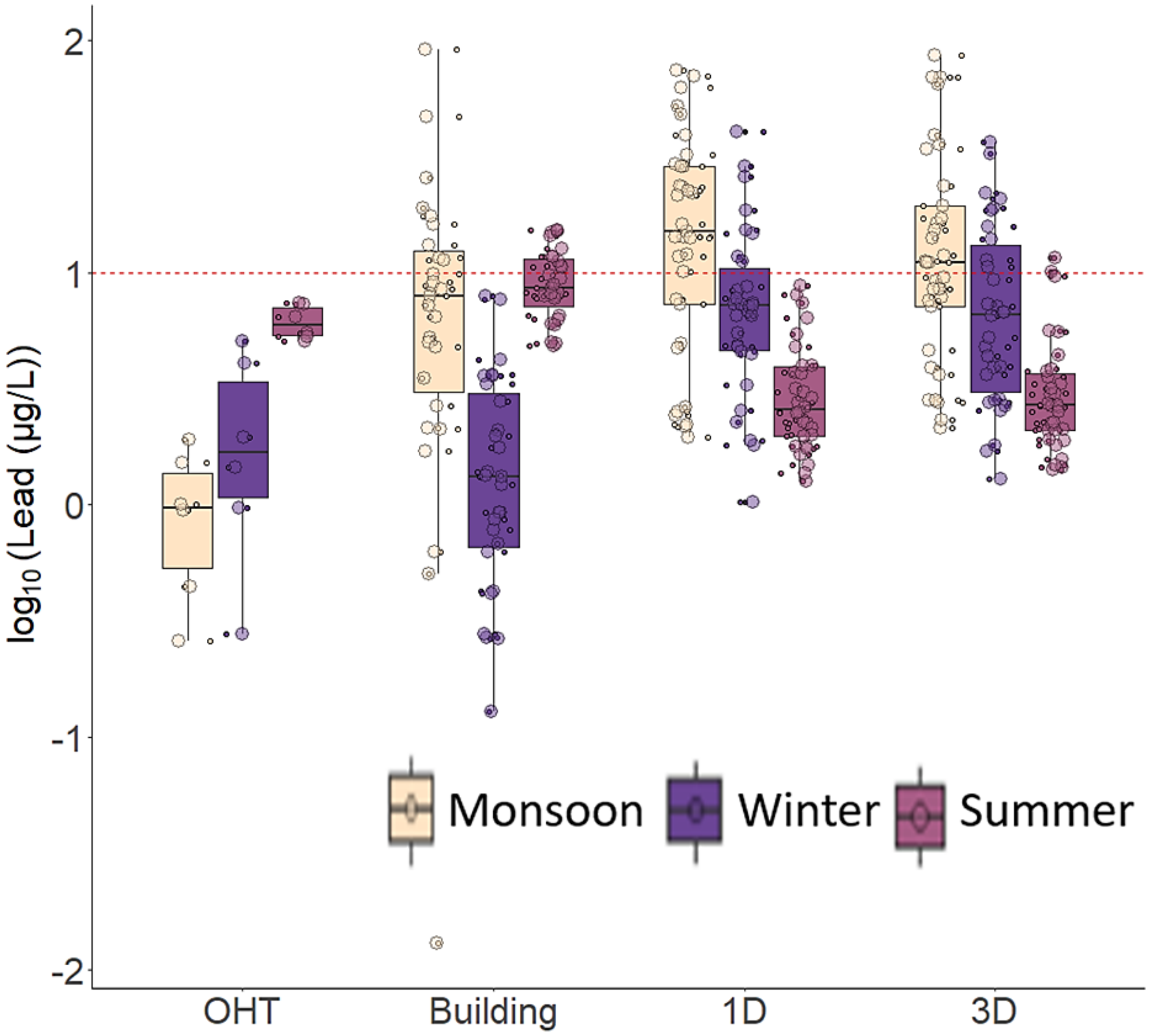


**Figure S9. Levels of Lead (µg/L) in tap water.** The y-axis represents the level of lead (µg/L) in the tap water collected from the OHTs and the buildings an year before the pandemic, and from the laboratory-scale water supply network in three different seasons (monsoon, winter, and summer) subject to water being drawn once per day (1D) and once in three days (3D) respectively. The Red dashed line represents the WHO (World Health Organisation, 2017) guidelines for heavy metal levels in drinking water, respectively.

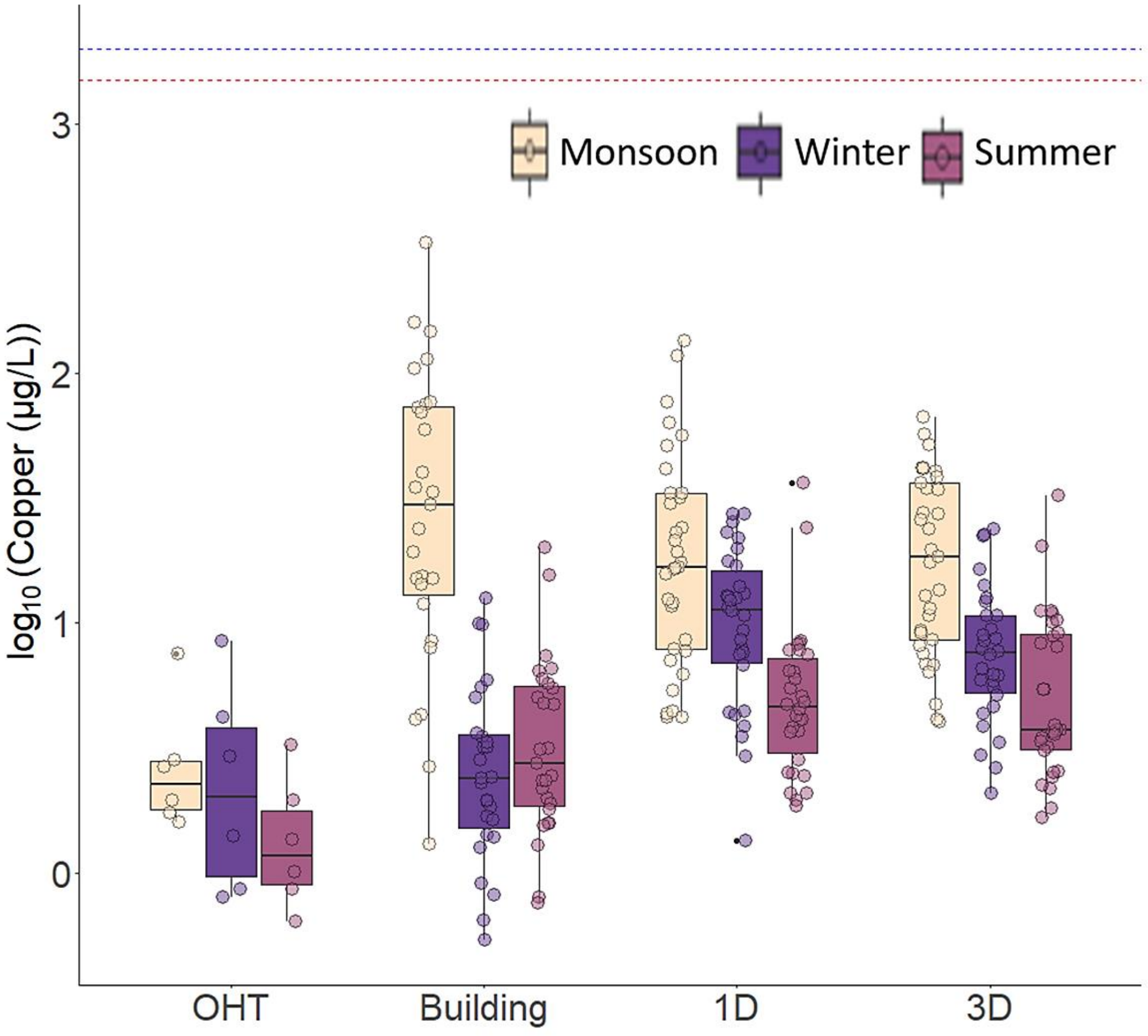


**Figure S10. Levels of Copper (µg/L) in tap water.** The y-axis represents the level of copper (µg/L) in the tap water collected from the OHTs and the buildings an year before the pandemic, and from the laboratory-scale water supply network in three different seasons (monsoon, winter, and summer) subject to water being drawn once per day (1D) and once in three days (3D) respectively. Red and blue dashed lines represent the WHO (World Health Organisation, 2017) and BIS (Bureau of Indian Standards, 2012) guidelines for heavy metal levels in drinking water, respectively.

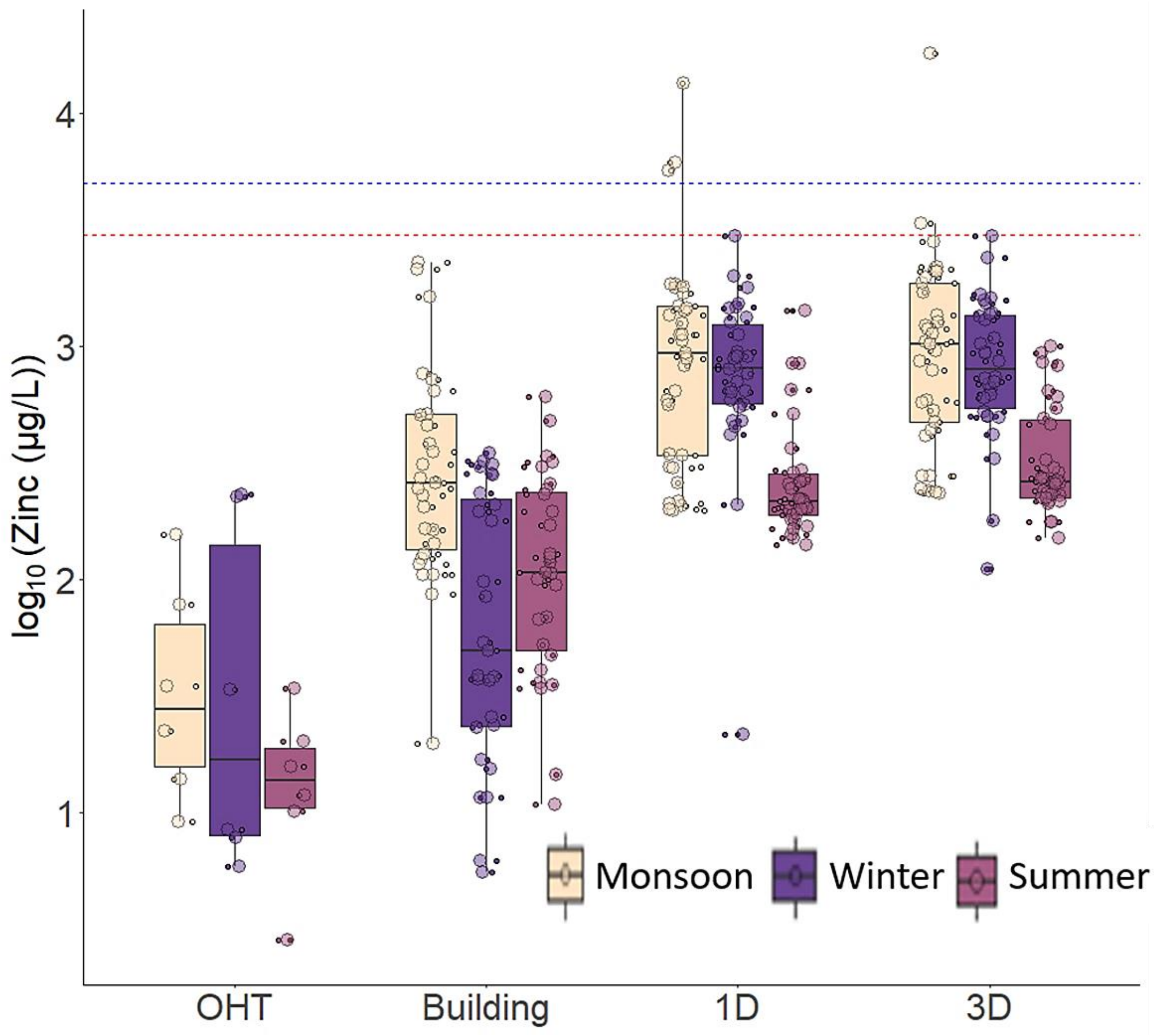


**Figure S11. Levels of Zinc (µg/L) in tap water.** The y-axis represents the level of zinc (µg/L) in the tap water collected from the OHTs and the buildings an year before the pandemic, and from the laboratory-scale water supply network in three different seasons (monsoon, winter, and summer) subject to water being drawn once per day (1D) and once in three days (3D) respectively. Red and blue dashed lines represent the WHO and BIS guidelines for heavy metal levels in drinking water, respectively.

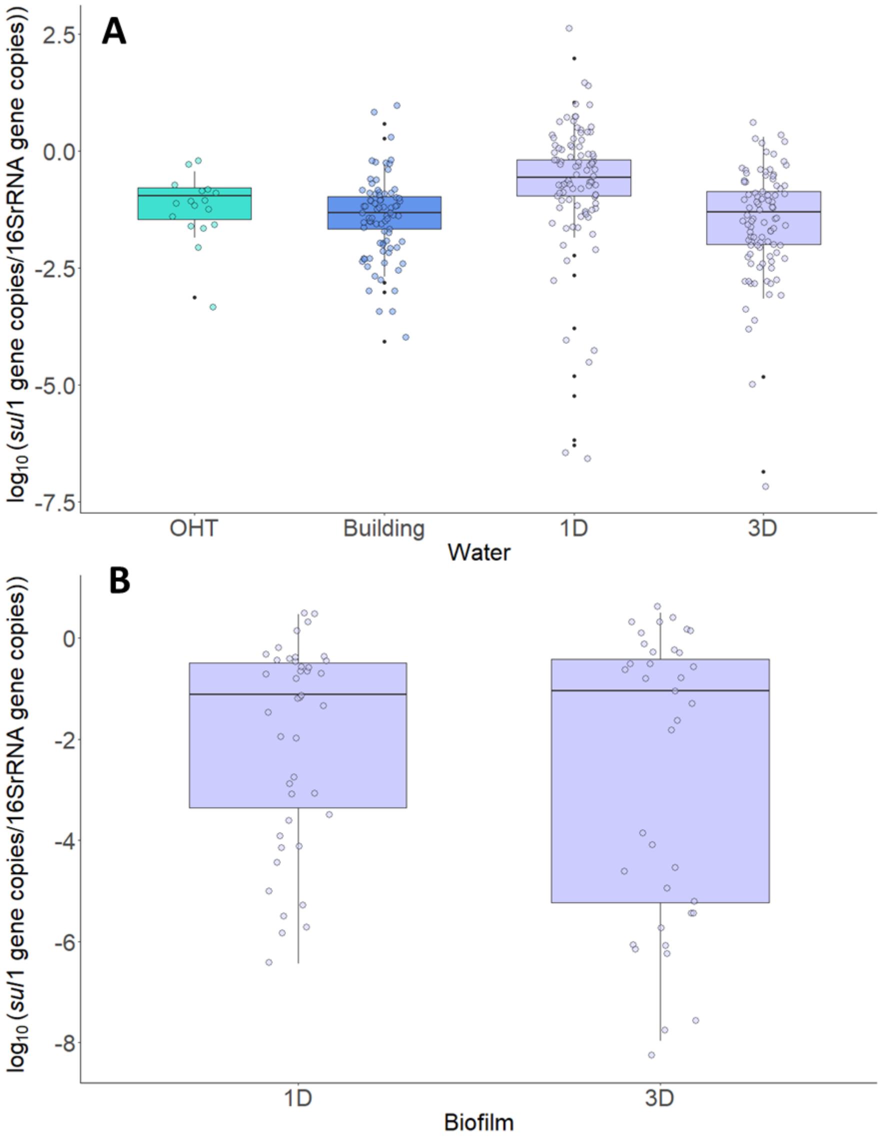


**Figure S13. Levels of *sul*1 gene per bacterium.** The y-axis represents the level of *sul*1 gene copies normalized to total bacterial count, assuming 4.2 16S rRNA gene copies per bacteria. **A.** The tap water and **B.** biofilm samples from the OHTs and the buildings an year before the pandemic, and from the laboratory-scale water supply network subject to water being drawn once per day (1D) and once in three days (3D) respectively.

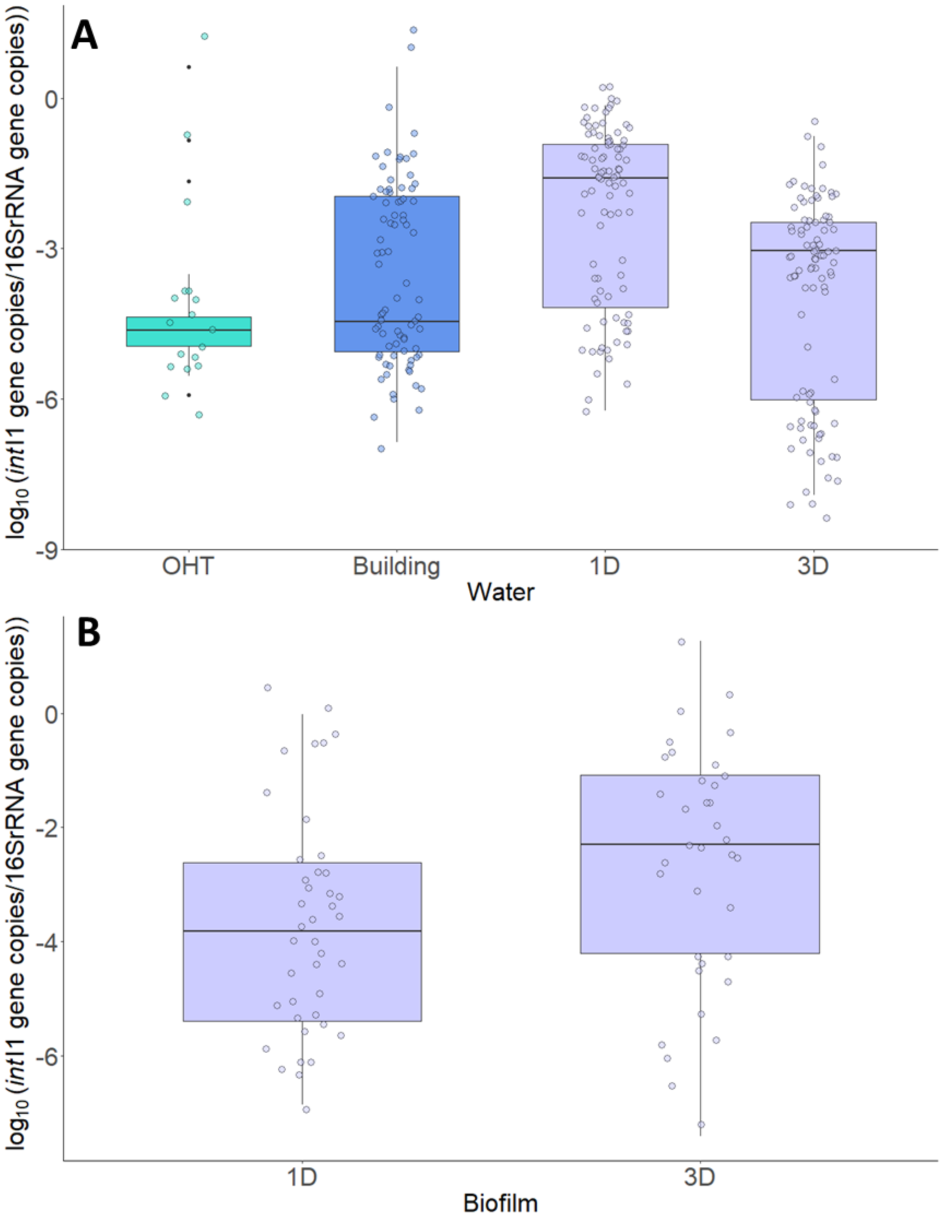


**Figure S15. Levels of *int*I1 gene per bacterium.** The y-axis represents the level of *int*I1 gene copies normalized to total bacterial count, assuming 4.2 16S rRNA gene copies per bacteria. **A.** The tap water and **B.** biofilm samples from the OHTs and the buildings an year before the pandemic, and from the laboratory-scale water supply network subject to water being drawn once per day (1D) and once in three days (3D) respectively.

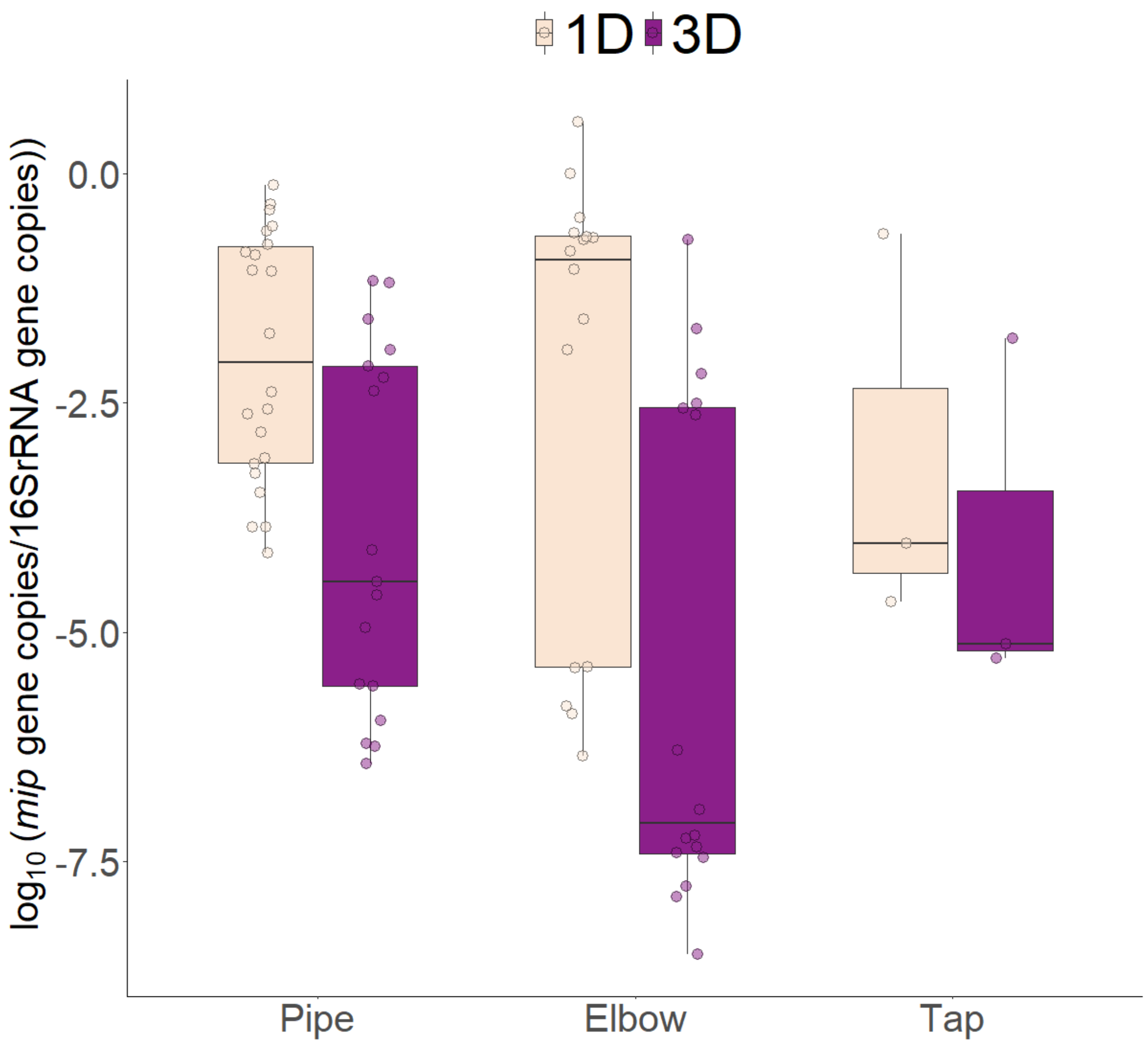


**Figure S16. Levels of *mip* gene per bacterium.** The y-axis represents the level of *mip* gene copies normalized to total bacterial count, assuming 4.2 16S rRNA gene copies per bacteria from biofilm samples swabbed from surfaces of pipes, elbows and taps subjected to different conditions: Prefixes 1D and 3D refer to water drawn once per day and once in three days respectively

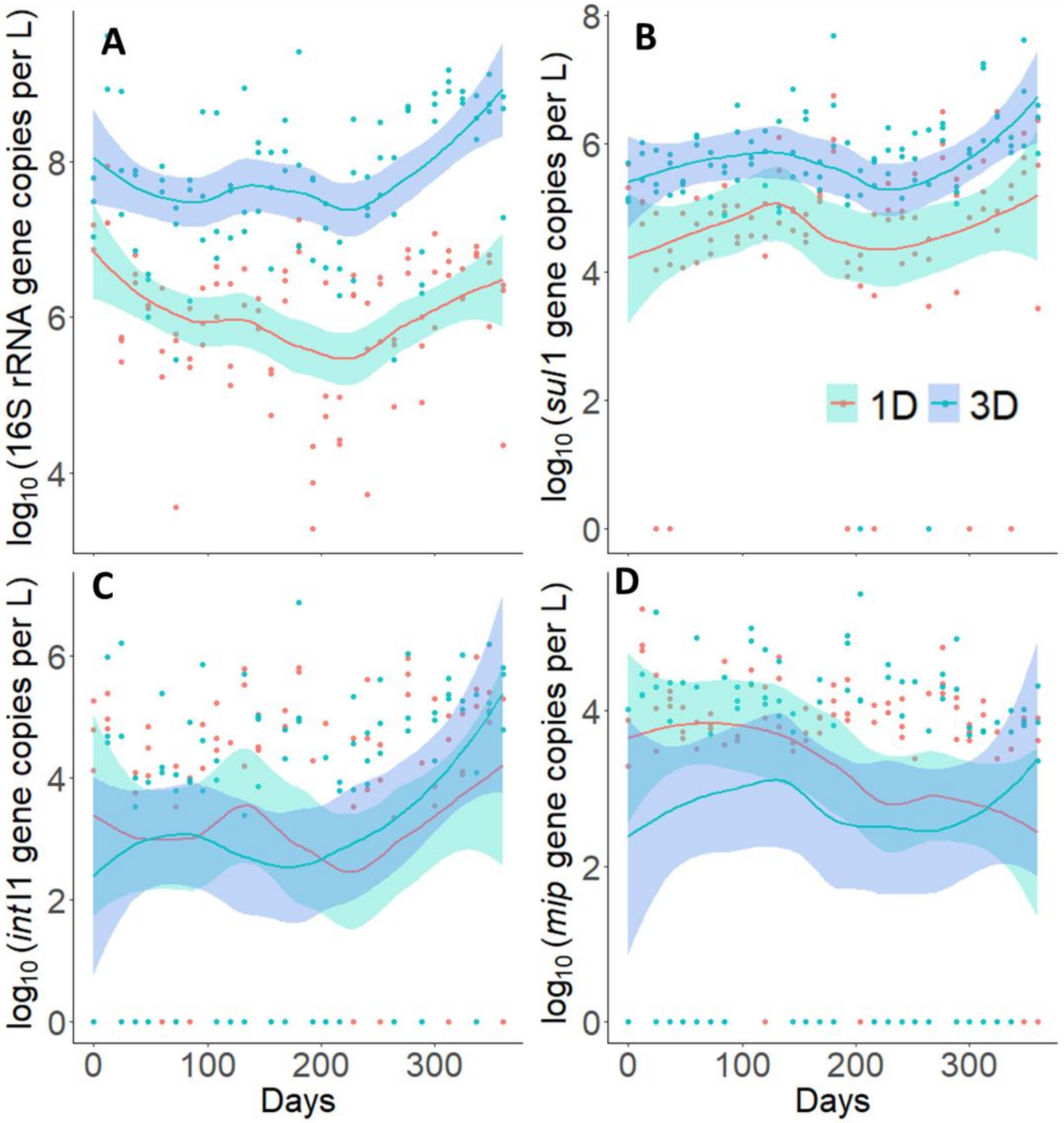


**Figure S18. Variation of 16S rRNA, *sul*1, *int*I1, *mip* genes variation of all genes throughout the year**

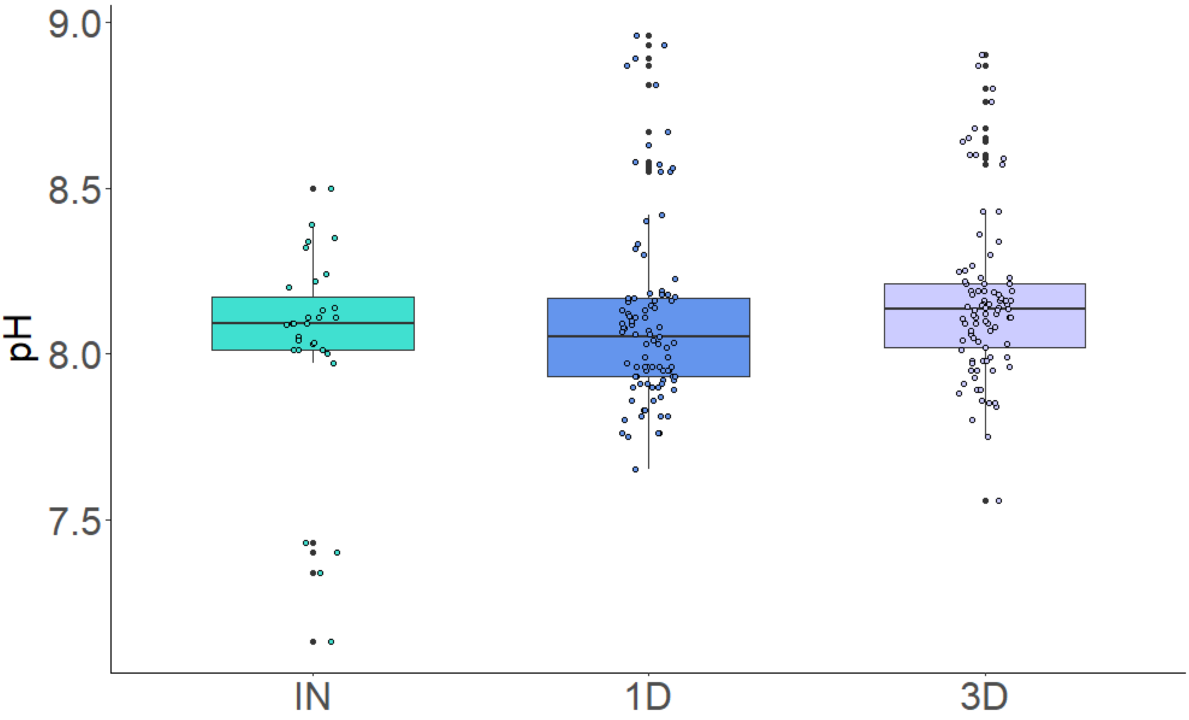


**Figure S18.** The pH values in water samples resolved across three different conditions (1D, 3D). IN represents the influent concentration.

**References:**


Bureau of Indian Standards, 2012. Indian Standard: Drinking water- Specification (second revision).

Chen, J., Yu, Z., Michel, F.C., Wittum, T., Morrison, M., 2007. Development and application of real-time PCR assays for quantification of erm genes conferring resistance to macrolides-lincosamides-streptogramin B in livestock manure and manure management systems. Appl. Environ. Microbiol. 73, 4407–4416. https://doi.org/10.1128/AEM.02799-06

Hembach, N., Schmid, F., Alexander, J., Hiller, C., Rogall, E.T., Schwartz, T., 2017. Occurrence of the mcr-1 colistin resistance gene and other clinically relevant antibiotic resistance genes in microbial populations at different municipal wastewater treatment plants in Germany. Front. Microbiol. 8, 1–11. https://doi.org/10.3389/fmicb.2017.01282

Lee, M.F., Peng, C.F., Hsu, H.J., Toh, H.S., 2011. Use of inverse PCR for analysis of class 1 integrons carrying an unusual 3′ conserved segment structure. Antimicrob. Agents Chemother. 55, 943–945. https://doi.org/10.1128/AAC.00988-10

Suzuki, M.T., Taylor, L.T., Long, E.F.D.E., 2000. Quantitative Analysis of Small-Subunit rRNA Genes in Mixed Microbial Populations via 5�-Nuclease Assays 66, 4605–4614.

Wajid, M., Saleemi, M.K., Sarwar, Y., Ali, A., 2019. Detection and characterization of multidrug-resistant Salmonella enterica serovar Infantis as an emerging threat in poultry farms of Faisalabad, Pakistan. J. Appl. Microbiol. 127, 248–261. https://doi.org/10.1111/jam.14282

Wang, H., Edwards, M., Falkinham, J.O., Pruden, A., 2012. Molecular survey of the occurrence of legionella spp., mycobacterium spp., pseudomonas aeruginosa, and amoeba hosts in two chloraminated drinking water distribution systems. Appl. Environ. Microbiol. 78, 6285–6294. https://doi.org/10.1128/AEM.01492-12

World Health Organisation, 2017. Guidelines for drinking-water quality, 4th edition: 1st addendum.
